# Identifying common disease trajectories of Alzheimer’s disease with electronic health records

**DOI:** 10.1101/2024.07.26.24311084

**Authors:** Mingzhou Fu, Timothy S. Chang

## Abstract

**Backgrounds:** Alzheimer’s disease (AD), a leading cause of dementia, poses a growing global public health challenge. While recent studies have identified AD risk factors, they often focus on specific comorbidities, neglecting the complex interrelations and temporal dynamics. Our study addresses this by analyzing AD progression through longitudinal trajectories, utilizing clinical diagnoses over time. Using machine learning and network analysis, we created a computational framework to identify common AD progression patterns.

**Methods:** We analyzed patient diagnoses from UC Health Data Warehouse’s Electronic Health Records, coded with the International Classification of Diseases, version 10 (ICD-10). Using the Fine and Gray model to detect significant temporal risk factors between diagnoses, we examined associations between diagnosis pairs and refined the patients’ diagnostic trajectories, delineating all possible trajectory combinations. These refined trajectories were compared using Dynamic Time Warping and grouped into clusters with hierarchical clustering. We investigated common AD trajectories through network analysis and compared patient demographics, symptoms, and AD manifestations across clusters. The Greedy Equivalence Search algorithm was used to infer causal relationships within these trajectories. We rigorously evaluated these trajectories through association tests and comparison to controls,

**Results:** Our analysis included 24,473 eligible AD patients, which was filtered to include 5,762 patients with 6,794 unique AD progression trajectories. We identified four trajectory clusters: 1) a mental health cluster (e.g., anxiety disorder → depressive episode) (N_patient = 1,448); 2) an encephalopathy cluster (e.g., hypertension → other disorders of brain) (N_patient = 3,223); 3) a neurodegenerative disease cluster (e.g., transient cerebral ischemic attacks → other degenerative disease of nervous system) (N_patient = 1,502); and 4) a vascular disease cluster (e.g. hypertension → other cerebrovascular diseases) (N_patient = 1,446). Significant differences were observed in demographics, symptoms, and AD features across clusters. Causal analysis indicated that 26.2% of the identified trajectory connections were causal. We also observed patients with risk trajectories faced higher risks of AD compared to those without the trajectory or with only a single risk factor.

**Conclusion:** We uncovered AD diagnosis trajectories, incorporating temporal aspects and causal relationships. These insights improve our understanding of AD development and AD subtypes, and can enhance risk assessment. Our findings can significantly benefit patient care and medical research by moving toward earlier and more accurate diagnoses, along with personalized treatment, such as medical risk factors management and lifestyle modifications.

## Background

Alzheimer’s disease (AD) is a progressive neurological disorder that mainly affects older adults, causing memory loss, cognitive decline, and difficulty with daily activities. Nearly 7 million Americans currently live with AD, and this number is expected to rise to almost 13 million by 2050. In 2021, Alzheimer’s was the fifth-leading cause of death among people aged 65 and older [1]. It is a significant public health concern, heavily burdening healthcare systems, caregivers, and families. Health and long-term care costs for people with dementia are projected to reach $360 billion in 2024 and nearly $1 trillion by 2050 [1].

Recent research on AD has identified several risk factors, such as cerebrovascular diseases [2,3], brain injury [4], depression [5], diabetes [6,7], and hearing loss [8]. However, most studies have focused on individual diseases and their direct links to AD, often ignoring the complex interactions and timing of these conditions. Understanding how these risk factors work together and in what order they occur is crucial for developing effective strategies to prevent and manage AD. Studying the disease trajectory, defined as a sequence of health events over time, can address this limitation. These events are ordered chronologically, often happen irregularly, and include diagnoses, medications, procedures, and lab tests [9]. Understanding these trajectories is crucial because it reveals the timing and order of potential risk factors, which can improve clinical decision-making and personalized care.

Current research on disease trajectories has several limitations. Much of the existing work builds on associated disease pairs, as in studies like Jensen et al. [10], which identify pairs of co-occurring sequential diseases with a statistically significant direction and combine these pairs into larger trajectories. Giannoula et al. [11] suggested a more straightforward approach involving matching diagnosis codes from patients’ records and extracting matched diagnosis sequences. Network-based frameworks are also used, identifying the shortest paths between diseases in the network, as shown by Dervić et al. [12]. While these methods have advanced our understanding, they often rely on model assumptions that oversimplify complex disease interactions and overlook important intermediate steps and nuances in disease progression. Moreover, there is often a lack of rigorous evaluation of the identified trajectories in these studies.

To address these gaps and provide a better understanding of AD progression, we developed a framework to identify common disease patterns from patients’ electronic health records (EHRs). We first used dynamic time warping (DTW) to align and compare the similarities between patients’ diagnosis sequences. Next, we applied unsupervised machine learning to cluster these diagnosis sequences and used network-based methods to identify the common trajectories within each cluster. Finally, we thoroughly evaluated these identified trajectories through association tests and comparing to controls. Our research contributes to the current literature by revealing the interconnected progression of various conditions leading to AD. By mapping these common trajectories, our findings can help predict disease progression and identify critical periods where timely medical or lifestyle interventions may significantly alter the course of the disease.

## Methods

The general study design is illustrated in **Figure 1**. The first step involves sample selection and EHR preprocessing to ensure the dataset is clean, standardized, and relevant for understanding AD trajectories. In the next step, we cleaned AD patients’ time-ordered diagnosis sequences by retaining connected diagnoses identified as significant patterns within a survival analysis framework. We then calculated the pairwise distances between these cleaned trajectories and clustered them into groups. Representative AD trajectories for each cluster were identified through network analyses. Finally, we conducted several analyses to evaluate the identified AD trajectory clusters. Detailed explanations for each step are provided in the following sections. All analyses were performed using R version 3.6 [13].

**Figure 1.**
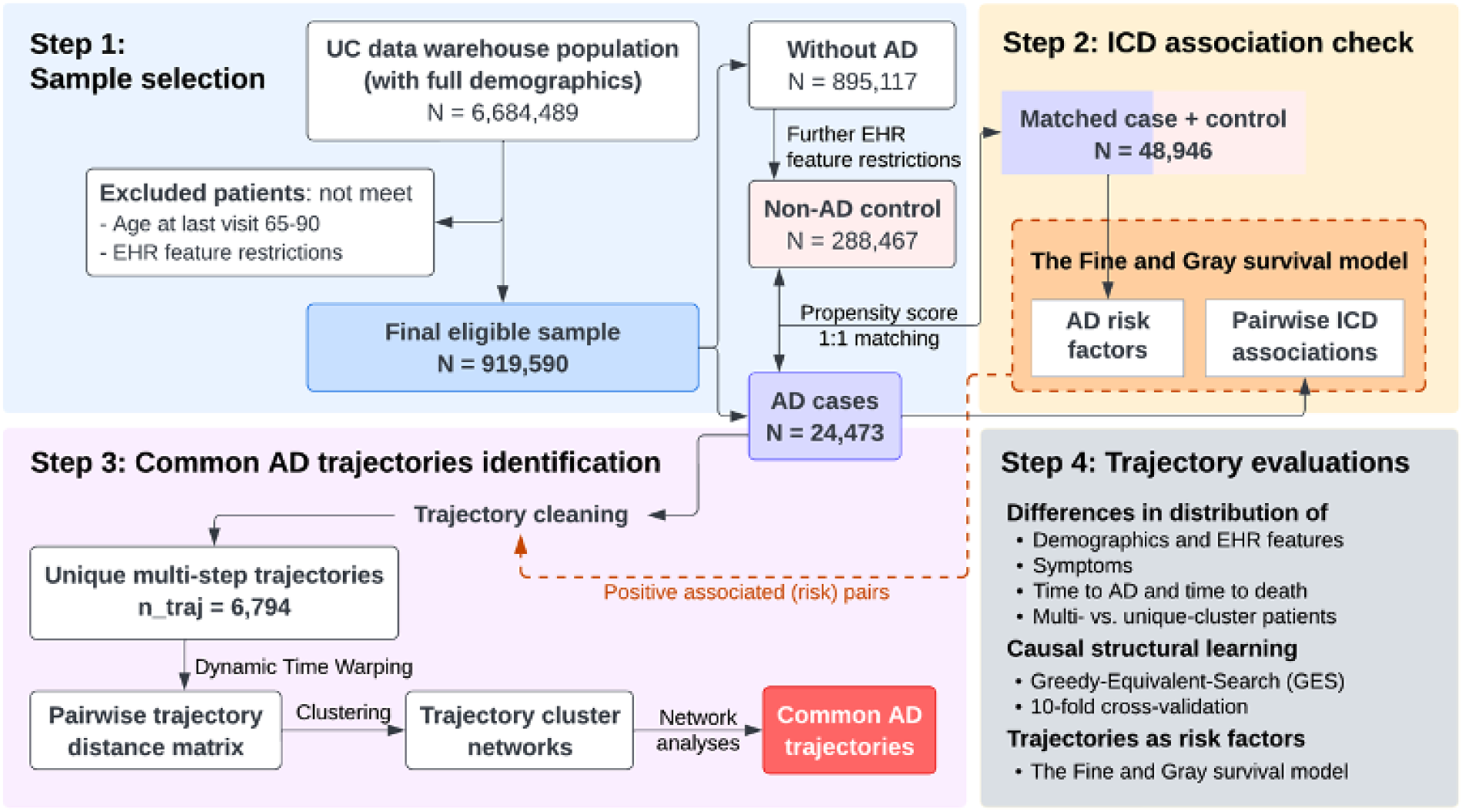
Study design.

### Sample selection in the UC Health Data Warehouse

#### Data source

The data for this study was extracted from the University of California Health Data Warehouse, a secure central EHR repository for the University of California Health System. This system includes 18 health professional schools, six medical centers, and ten hospitals, and it encompasses data on 8.7 million patients seen since 2012. The data is de-identified to facilitate clinical research under the guidance of institutional review boards, privacy and compliance officers, and information security officers [14].

#### EHR preprocessing

We first ensured the accuracy and completeness of the data by conducting patient-level cleaning. For each patient, we calculated key EHR features, including the length (in years) and density (encounters per year) of their medical records. Patients were included if they met the following criteria: complete records with no missing demographics (age, sex, race/ethnicity), at least two encounters on different dates with at least one encounter per year, and aged between 65 and 90 at their last encounter. This age range was chosen to include patients likely to develop AD, with an upper limit set because the EHR dataset censors patients at this age [15].

At the record level, we truncated the International Classification of Diseases, 10th Revision (ICD-10) codes to their first three digits to standardize diagnoses to phenotypes of appropriate granularity [16]. The date of the first encounter with each three-digit ICD code was recorded as the diagnosis date. ICD codes from Chapters XV (Pregnancy, childbirth, and the puerperium), XVI (Certain conditions originating in the perinatal period), XVII (Congenital malformations, deformations, and chromosomal abnormalities), and XX (External causes of morbidity and mortality) were removed as they are less likely to be associated with an individual’s AD onset. In addition, ICD codes from Chapter XVIII (Symptoms, signs, and abnormal clinical and laboratory findings, not elsewhere classified) were excluded from the primary identification of disease trajectories as these are primary symptoms but were retained for downstream analyses.

#### Case Definition

Patients were identified as having AD if they had at least one encounter with an AD ICD diagnosis code (G30). The date of the first encounter with this diagnosis was considered the AD diagnosis date. Only diagnoses occurring on or before this date were retained for analyzing disease trajectories.

### Pairwise ICD codes association check in a sampled population

We used a survival analysis framework to identify risk-associated diagnoses in patients by evaluating the association between pairwise ICD codes. This step was taken to simplify AD patients’ trajectories (see details later in *Trajectory Cleaning and backward-building*).

#### Sample Set

To identify AD temporally associated risk factors, we selected all AD cases and matched non-AD controls as the discovery sample. For eligible non-AD controls, we applied stricter EHR feature criteria to minimize underdiagnosis and misclassification of potential AD. These criteria included a record length of at least five years, a minimum of two encounters, and a record density of at least two encounters per year. We matched controls to AD cases using propensity score matching [17] based on age at the last visit, gender, race/ethnicity, and location sources, with a matching ratio of 1:1. For other pairwise ICD code associations, we tested within AD cases only, interpreting these as significant associations within the AD subpopulation.

#### Fine and Gray model

We used the Fine and Gray (FG) proportional subdistribution hazards model [18] for our analysis. This model handles competing risks, such as death, by keeping them in the risk set indefinitely [18]. We calculated the follow-up time for each patient, considering both the time to event and censorship. Follow-up time can differ for the same individual depending on the specific outcome of interest. Exposure ICD codes were treated as binary variables, indicating whether the condition was present before the outcome. We included age, sex, race/ethnicity, and record length as predictor variables to control for patient baseline differences. We identified positive risk patterns using an adjusted p-value of ≤ 0.1 (False Discovery Rate for multiple testing [19]) and a Hazard Ratio (HR) > 1.

### Patient trajectories preprocessing

#### Trajectory cleaning and backward-building

During preprocessing, diagnoses occurring at the same time as an AD diagnosis were moved to one step before the AD diagnosis. This ensured the retention of important diagnostic patterns, contributing to longer and more informative trajectories. We then identified all possible trajectories for patients with three or more steps, splitting them into multiple sequences and greedily concatenating them to form longer trajectories (**Supplementary Figure 1**). The backward-building approach started from the AD diagnosis, with the step preceding AD being an AD risk factor identified from the FG models. Only positively associated ICD pairs were connected, retaining only risk patterns in the diagnostic sequence and improving interpretability of longer disease trajectories For each diagnosis, we considered every possible connection in preceding nodes and split trajectories for multiple diagnoses at the same step. Greedy concatenation prioritized longer trajectories, retaining only those with three or more steps.

#### Two-step-trajectory expansion

To retain a significant portion of the AD population, we expanded the trajectories of patients with only two encounters (N = 6,265, approximately 25.6% of the total) to three-step trajectories using risk pairs from the FG models. We assumed that diseases could progress in a specific order, even if diagnoses occurred on the same visit date. An unweighted directed network based on significant pairs was constructed, starting from all co-occurring diagnoses leading to G30. This process expanded the records from two encounters to all possible three-step paths, adding 1,571 patients with expanded three-step trajectories.

#### Sampling of trajectories

In the final step, we adjusted extremely long trajectories. The number of steps in the cleaned trajectories ranged from three to 17. To minimize bias in the DTW alignment step (see details later in *Calculate distances between trajectories)*, the maximum number of steps was set to nine, covering 99% of patients. For the 1% (N = 36) with trajectories longer than nine steps, we identified their nine-step trajectories by loosening greedy concatenation restrictions. Each patient was limited to a maximum of three trajectories, the median for eligible patients. For patients with more than three trajectories (N = 2,846), we performed sampling by distances to retain distinct trajectories. A DTW distance matrix was built for each patient, clustering based on distance with three clusters, and one trajectory was sampled from each cluster.

### Common AD trajectory identification

#### Calculate distances between trajectories

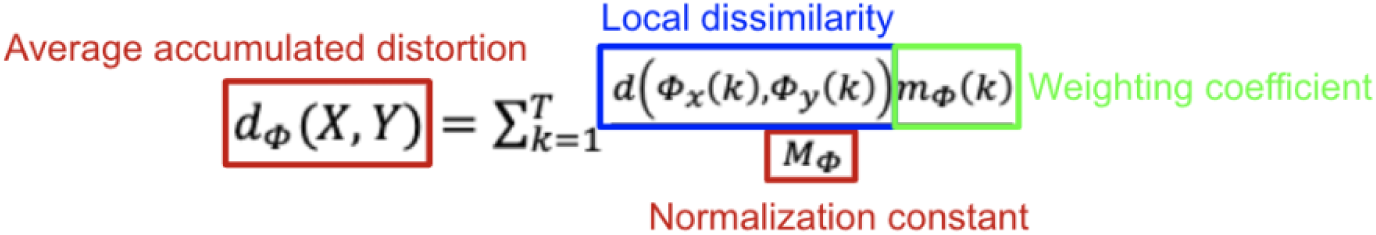

We used the DTW [20] algorithm to align and calculate distances between patient trajectories. DTW has advantages over Euclidean distance, particularly in handling sequences of varying lengths and temporal distortions. The core component of DTW is the average accumulated distortion, which is the sum of distances minimized during alignment [20]. Local dissimilarity measures the distance between two ICD diagnoses in each time series, which was determined based on the Systemized Nomenclature of Medicine, Clinical Terms (SNOMED) embedding similarities established in our previous work [21]. We applied the asymmetric P05 step pattern to align trajectories, with slope weights to each step. The Open-Begin-End Dynamic Time Warping (OBE-DTW) method was used to allow for partial alignment with free endpoints, enabling the skipping of longer segments [22]. A normalization constant ensured that accumulated distortions were comparable across paths of different lengths. Finally, we obtained a trajectory distance matrix with dimensions 6794 by 6794, representing the pairwise distances between unique patient trajectories.

#### Trajectory clustering

Next, we clustered the trajectory distance matrix. We tested k-means [23], hierarchical [24], and partition around medoids (PAM) [25] clustering methods. To determine the optimal number of clusters and the best clustering algorithm, we evaluated them using several indices: the

Calinski–Harabasz (CH) index, where higher values indicate better clustering; the Davies– Bouldin (DB) index, where lower values are preferable; and the Silhouette (Sil) index, where higher values indicate better-defined clusters [26]. To ensure the robustness of our clustering results, we conducted a sensitivity analysis using the adjusted rand index, which measures the similarity between different clustering outcomes [27].

#### Network analyses

To identify common AD trajectories in each cluster, we first built raw networks using patient trajectories, focusing on connections followed by at least 0.5% of patients, particularly paths to AD (G30). We then extracted the backbones of each cluster network using modularity vitality, which measures the significance of nodes and edges based on their contribution to modularity [28]. Higher modularity indicates a stronger community structure. We pruned the network by removing nodes and edges with the lowest contribution to the network structure one at a time, balancing simplicity with higher network modularity, and retaining only the most critical ones for the community structure. We ensured all paths to AD were retained in the simplified network by taking the intersection of remaining nodes and edges. The node with the highest contribution was identified as the central node for each cluster. We reported the top five trajectories shared by AD patients within each cluster as the typical trajectory and also examined the decomposition of central nodes with more than three digits.

### Evaluation of common AD trajectories

#### Patient characteristics across clusters

After data cleaning, patients can have up to three unique trajectories, meaning they may belong to different clusters. We first examined the distribution of patients across clusters using a Venn diagram [29]. To compare patient characteristics across clusters, we excluded those who fell into more than one cluster (N = 463) from the primary analysis. However, we included all patients for sensitivity analysis to check for consistency.

Distributions of demographic and EHR features were compared across clusters. ANOVA [30] and Kruskal-Wallis tests [31] were used to determine statistical significance. Additionally, we examined the distributions of symptoms (Chapter XVIII: Symptoms, signs, and abnormal clinical and laboratory findings) across clusters. We focused on symptoms related to cognition, perception, emotional state, and behavior at the three-digit ICD code level, as these are most relevant to AD. Symptoms from other systems were grouped into subchapters (two-digit ICD codes) for easier comparison. We analyzed symptoms occurring at all times, three years before, and one year before the first AD diagnosis. We also compared the time elapsed from the central node to AD for each cluster. Boxplots were used to visualize results, and the Wilcoxon Rank-Sum test [32] was employed for pairwise comparisons.

We compared AD features across different clusters, including age of onset, time from the first AD diagnosis to the most recent visit, and time to death (if applicable). Kaplan-Meier curves [33] were used to compare time from the first visit to the first AD diagnosis and from the first AD diagnosis to death. The log-rank test [34] was used to determine the statistical significance of survival curves. Finally, we examined the distributions of other ICD codes, comparing alive and deceased patients within each cluster to identify potential risk factors leading to death within a cluster.

#### Causal structural learning

To determine the causality of the identified trajectories, we used the Greedy Equivalent Search (GES) algorithm [35] to learn the causal structure of AD from our EHR sample. GES is a score-based method that starts with an empty, partially Directed Acyclic Graph (DAG) and greedily adds and removes edges to maximize the score, such as the Bayesian Information Criterion (BIC). This approach is computationally efficient providing insights into possible causal structures [36].

We used the same 1:1 matched case-control sample from the FG modeling to learn the causal structure. To ensure the robustness of our results, we applied a 10-fold cross-validation approach. In each iteration, we learned the DAG with all the data, leaving one-fold out each time, and repeated this for ten iterations. We only selected causal edges that appeared in at least five of the ten iterations, ensuring consistent and reliable identification of causal relationships.

#### Risk trajectories testing in AD and controls

To confirm that the identified disease trajectories are real risk factors for AD, we tested the associations between these risk trajectories and the incidence of AD in a newly sampled population, treating the risk trajectory as a single binary variable. We redefined our control group to include only patients who have never been diagnosed with AD or any exclusive phenotypes previously defined [37]. We examined these associations using a similar FG proportional subdistribution hazards model framework. Two models were tested: one comparing those with the risk trajectory to all others, and another comparing those with the risk trajectory to those with any of the diagnoses in the risk trajectory.

## Results

### Sample description

From the initial dataset, 919,590 patients from the University of California Health Data Warehouse met the inclusion criteria and were included in the study. Among these eligible patients, 24,473 (2.7%) had at least one documented AD diagnosis based on the ICD-10 code definition.

In all AD patients, we examined co-occurring diagnoses with AD at the full-digit ICD level. We found that 73% of AD patients also had an F02.8 diagnosis (Dementia in other diseases classified elsewhere) at the same visit as their AD diagnosis, and 95% had an F02.8 diagnosis within 90 days before their AD diagnosis. This suggests that F02.8 is often a co-diagnosed given at the same time as AD [38] and may be due to co-mapping based on EHR medical vocabularies [39]. For example, G30 and F02.80 will both be mapped when “Alzheimer’s disease” is search and selected as a diagnosis codes. To avoid confusion in later trajectory analysis, we removed F02.8 diagnoses occurring within 90 days before the AD diagnosis but kept other instances of F02.8.

### Pairwise ICD codes temporal associations

Firstly, we used all AD cases and 1:1 matched non-AD controls to identify AD temporal risk factors. After matching based on age at the last visit, gender, race/ethnicity, and location sources, there were no significant differences in demographic and EHR feature distributions between cases and controls (**Supplementary Table 1**).

In the AD cases and matched non-AD control sample (N = 48,946), the FG results identified 16 ICD codes positively associated with the incidence of AD after adjusting for the false discovery rate, suggesting potential risk factors of AD (**Table 1**). Most of these risk factors were from the ICD Chapter on mental, behavioral, and neurodevelopmental disorders. Among these, unspecified dementia (F03) had the highest HR, indicating that patients with unspecified dementia have a 3.45-fold increased risk of developing AD compared to controls, even when accounting for the competing risk of death. Other risk factors included various mental, behavioral, neurodevelopmental disorders, and diseases of the nervous system.

**Table 1.** Risk factors of Alzheimer’s disease identified by the Fine and Gray proportional subdistribution hazards model, UC sample (N = 48,946)

| ICD | HR | Adjusted P | Description | Chapter |
| --- | --- | --- | --- | --- |
| F03 | 3.45 | <0.001* | Unspecified dementia | Mental, Behavioral and<br>Neurodevelopmental disorders |
| F09 | 2.39 | <0.001* | Unsp mental disorder due to known physiological condition |  |
| F01 | 2.30 | <0.001* | Vascular dementia |  |
| F07 | 2.29 | <0.001* | Personality & behavrl disorders due to known physiol cond |  |
| G31 | 2.27 | <0.001* | Oth degenerative diseases of nervous system, NEC | Diseases of the nervous system |
| F29 | 1.91 | <0.001* | Unsp psychosis not due to a substance or known physiol cond | Mental, Behavioral and<br>Neurodevelopmental disorders |
| F02 | 1.89 | <0.001* | Dementia in other diseases classified elsewhere |  |
| F22 | 1.71 | <0.001* | Delusional disorders |  |
| F06 | 1.53 | <0.001* | Other mental disorders due to known physiological condition |  |
| F31 | 1.50 | <0.001* | Bipolar disorder |  |
| G91 | 1.47 | <0.001* | Hydrocephalus | Diseases of the nervous system |
| I67 | 1.30 | <0.001* | Other cerebrovascular diseases | Diseases of the circulatory system |
| F39 | 1.18 | 0.003 | Unspecified mood [affective] disorder | Mental, Behavioral and<br>Neurodevelopmental disorders |
| F10 | 1.17 | <0.001* | Alcohol related disorders |  |
| F32 | 1.10 | <0.001* | Major depressive disorder, single episode |  |
| G93 | 1.10 | <0.001* | Other disorders of brain | Diseases of the nervous system |

For other pairwise ICD code associations, we tested within AD cases (N = 24,473) only, focusing on ICDs with a prevalence of over 1% in the AD population (N_ICD = 302, excluding AD). We performed association tests using the FG model between these pairwise ICD codes. After adjusting for the false discovery rate, we found 32,654 pairs with significant positive associations, suggesting potential risky patterns in the diagnostic sequence. These risky patterns were used in the trajectory cleaning steps.

### Common AD trajectories identification

#### DTW alignment and clustering

We started with the full cohort of AD patients (N = 24,473) from the University of California Health Data Warehouse to build AD trajectories. After trajectory cleaning and backward-building, 5,762 AD patients with 6,794 unique 3-9 step trajectories were used for DTW alignment, among which 1,571 (27%) patients had expanded 3-step trajectories. The average number of steps in each trajectory was six, and each patient had an average of 2.3 trajectories after cleaning.

We used the DTW algorithm [20] to align and calculate distances between patient trajectories. This produced a trajectory distance matrix with dimensions of 6794 by 6794, representing the pairwise distances between unique patient trajectories. Based on evaluation metrics, we selected k-means clustering with four clusters (**Supplementary Figure 2**). Sensitivity analyses showed moderate agreement between the different clustering methods, with adjusted rand indices of 0.13 between k-means and hierarchical clustering, 0.20 between k-means and PAM, and 0.23 between PAM and hierarchical clustering.

#### Trajectory cluster characteristics

We identified four clusters from the AD patients, each with unique characteristics and some shared diagnoses and paths. **Supplementary Figure 3** shows the raw networks of each cluster using patient trajectories followed by at least 0.5% of patients. Simplified networks after backbone extraction are shown in **Figures 2A-D**.

**Figure 2A.**
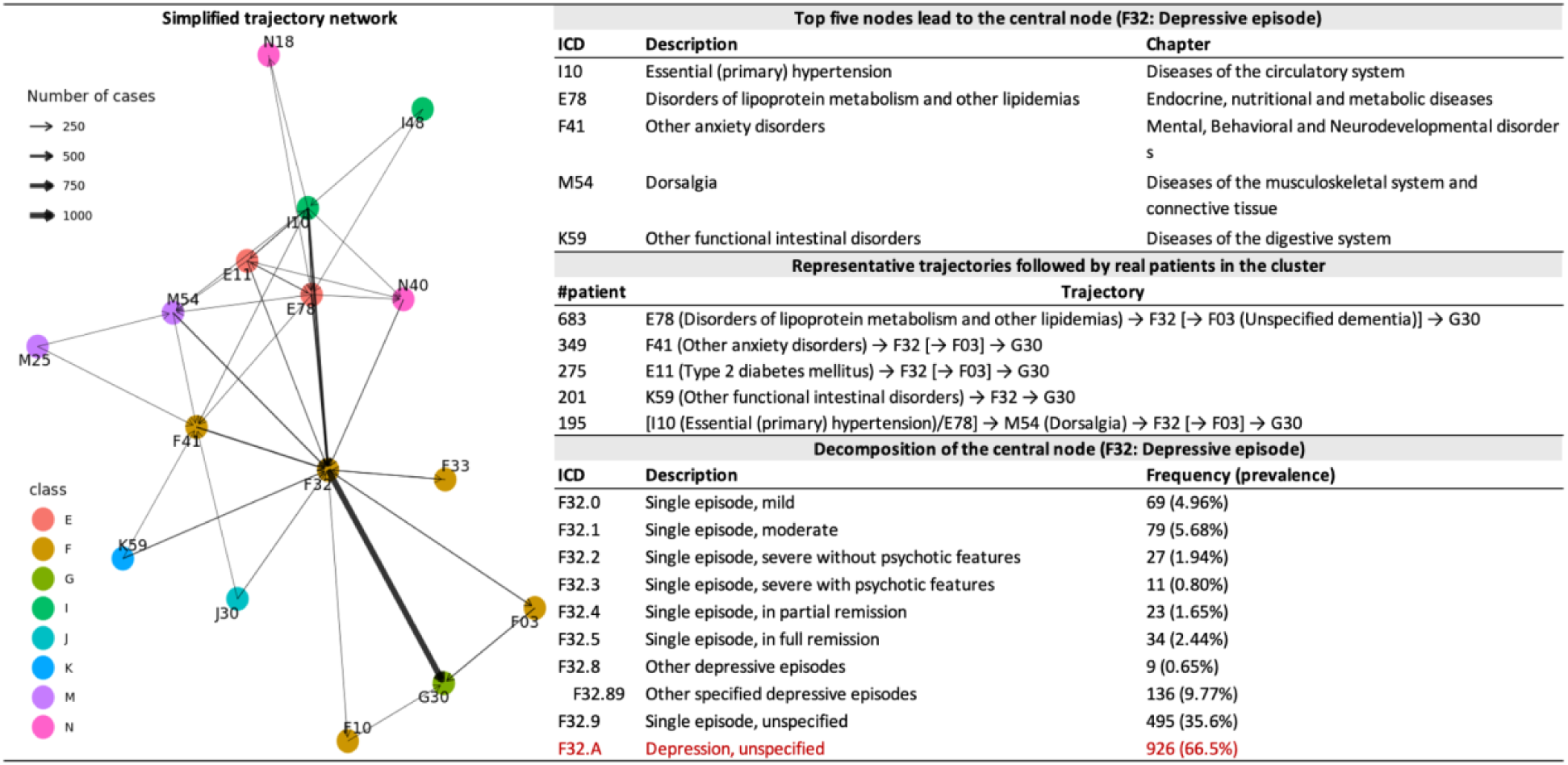
Summary of the depression AD trajectory (N_patient = 1,448, N_traj = 1,266). Decomposition prevalence not adding up to 100% - one patient could have multiple diagnoses.

The first cluster centers around mental illness, with depressive episode (F32) as the central node (**Figure 2A**). It includes 1,448 patients with 1,266 unique diagnosis trajectories. Common preceding diagnoses leading to F32 are circulatory system diseases like essential hypertension (I10) and atrial fibrillation (I48), metabolic diseases such as disorders of lipoprotein metabolism (E78) and type 2 diabetes (E11), mental disorders like anxiety disorders (F41), musculoskeletal diseases such as dorsalgia (M54), and digestive system diseases like other functional intestinal disorders (K59). Representative trajectories include E78 → F32 → G30 and F41 → F32 → G30. Notably, 67% of patients in this cluster were diagnosed with unspecified depression (F32.A).

The second cluster focuses on brain diseases, with other disorders of the brain (G93) as the central node, involving 3,223 patients and 764 unique trajectories (**Figure 2B**). The most prevalent full G93 ICD code diagnosis within this cluster encephalopathy, unspecified G93.40 (45%). Preceding diagnoses include circulatory system diseases like essential hypertension (I10) and other cerebrovascular diseases (I67), and genitourinary system diseases such as other diseases of the urinary system (N39) and benign prostatic hyperplasia (N40). Representative trajectories include I10 → G93 → [I67] → G30 and E78 → G93 → G30.

**Figure 2B.**
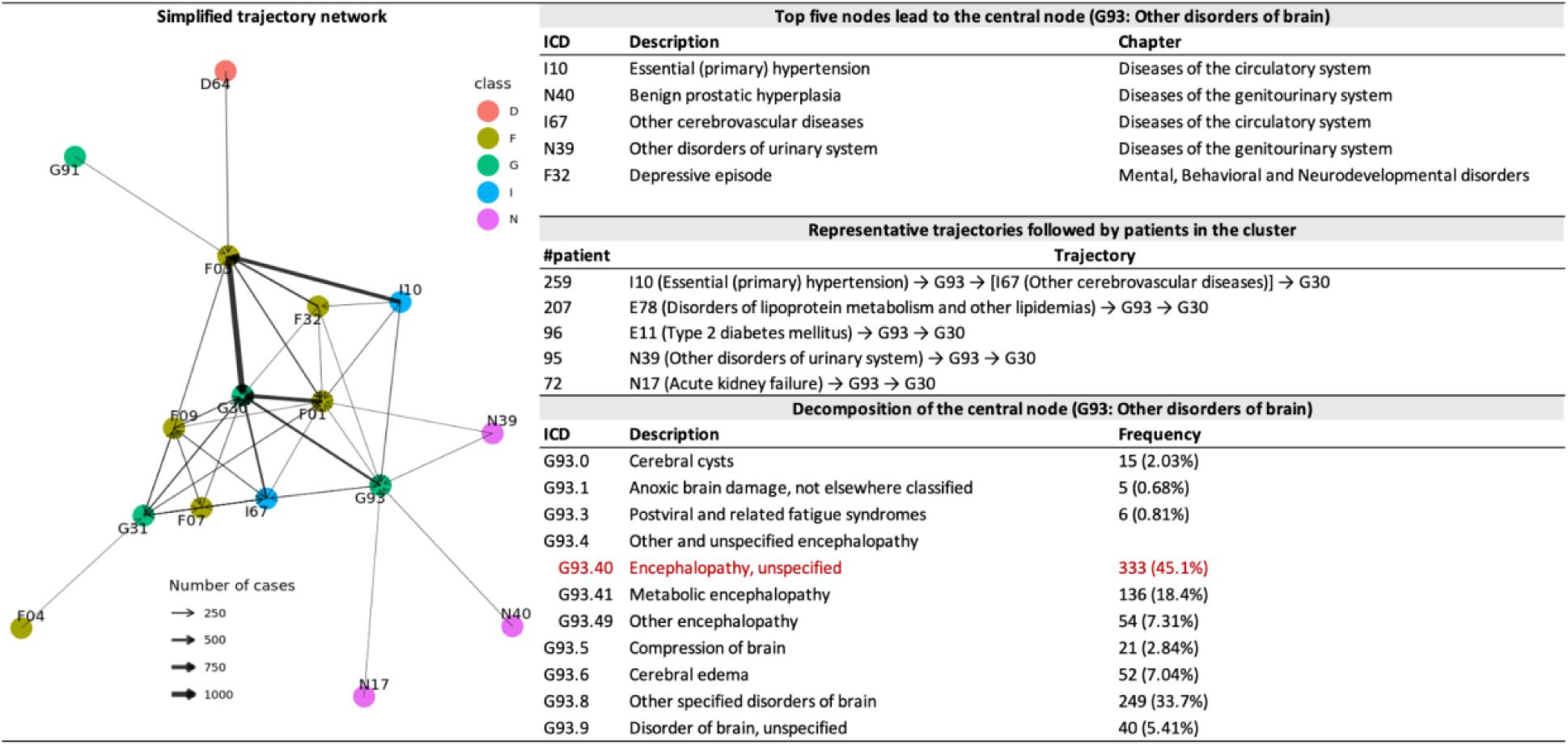
Summary of the encephalopathy disease AD trajectory (N_patient = 3,223, N_traj = 764)

The third cluster deals with neurodegenerative diseases, with other degenerative diseases of the nervous system (G31) as the central node, comprising 1,502 patients and 1,658 unique trajectories (**Figure 2C**). In this cluster, 70% of patients had mild cognitive impairment (MCI) of uncertain or unknown etiology (G31.84), a full ICD code for G31. Prior diagnoses often include nervous system diseases like transient cerebral ischemic attacks (G45), genitourinary system diseases such as menopausal disorders (N95) and male erectile dysfunction (N52), and ear and mastoid process diseases such as other inflammation of eyelid (H01) and disorders of the lacrimal system (H04). Prominent trajectories are G31 → F01 (vascular dementia) → G30, H04 → G31 → G30, and G45 → G31 → G30.

**Figure 2C.**
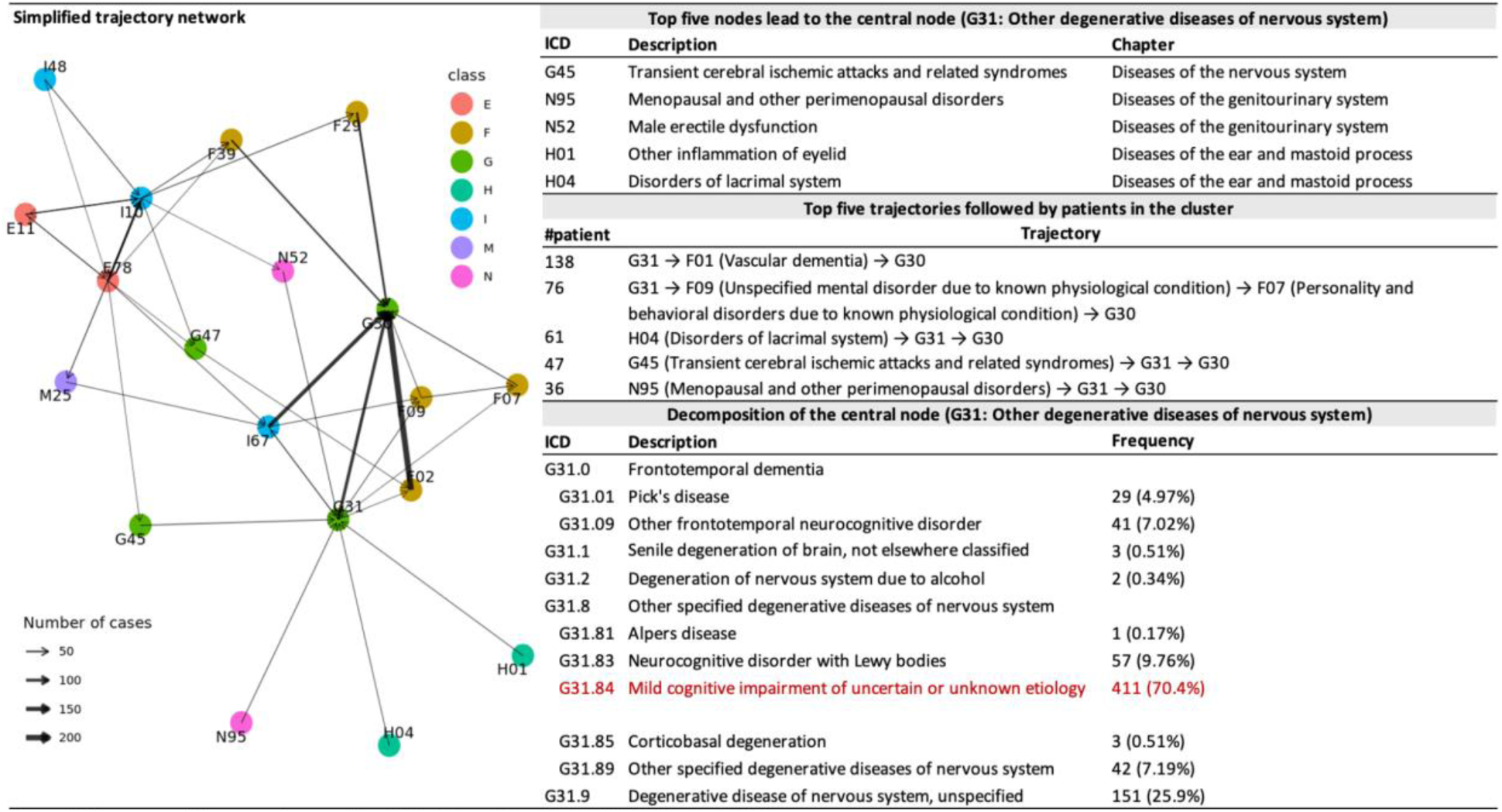
Summary of the neurodegeneration AD trajectory (N_patient = 1,502, N_traj = 1,658)

The final cluster centers on vascular diseases, with other cerebrovascular diseases (I67) as the central node, including 1,446 patients and 3,106 unique trajectories (**Figure 2D**). Common preceding diagnoses are musculoskeletal diseases like other joint disorders (M25), unspecified soft tissue disorders (M79), and dorsalgia (M54), and conditions caused by postprocedural states (Z98). Notable trajectories are E78 → I10 → I67 → G30, I10 → D64 (other anemias) → F03 → G30, and M25 → I67 → G30. In this group, 44% of patients had unspecified cerebrovascular disease (I67.9).

**Figure 2D.**
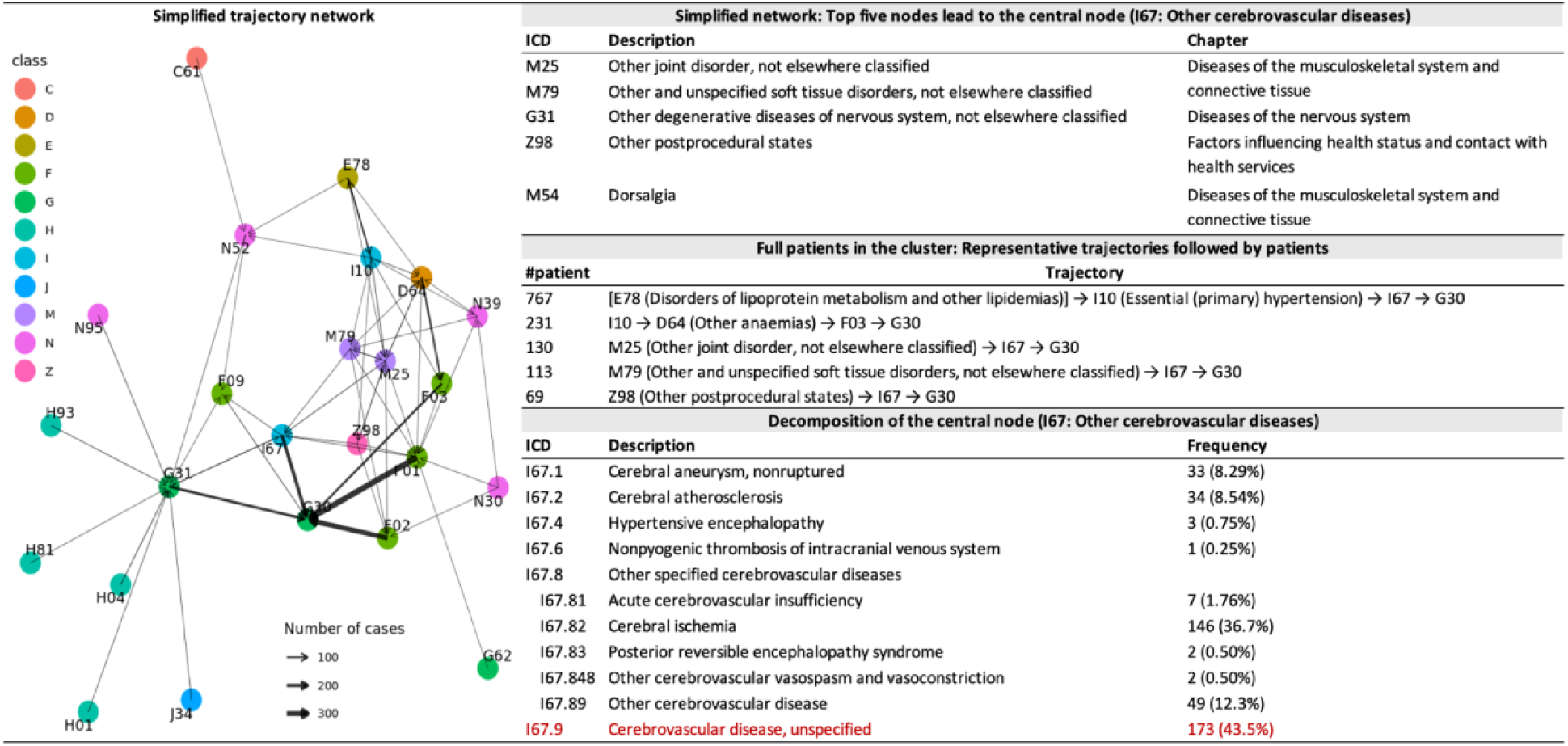
Summary of the vascular disease AD trajectory (N_patient = 1,446, N_traj = 3,106)

### Evaluation of common AD trajectories

#### Patient characteristics across clusters

We analyzed the distribution of patients across clusters. Out of the total, 4,078 patients fell into only one cluster, 1,511 patients into two clusters, and 173 patients into three clusters. **Figure 3** illustrates the patient distribution with a Venn diagram. Notably, the MCI cluster tends to overlap with other clusters, indicating that AD patients often have comorbidities in addition to cognitive impairment trajectories.

**Figure 3.**
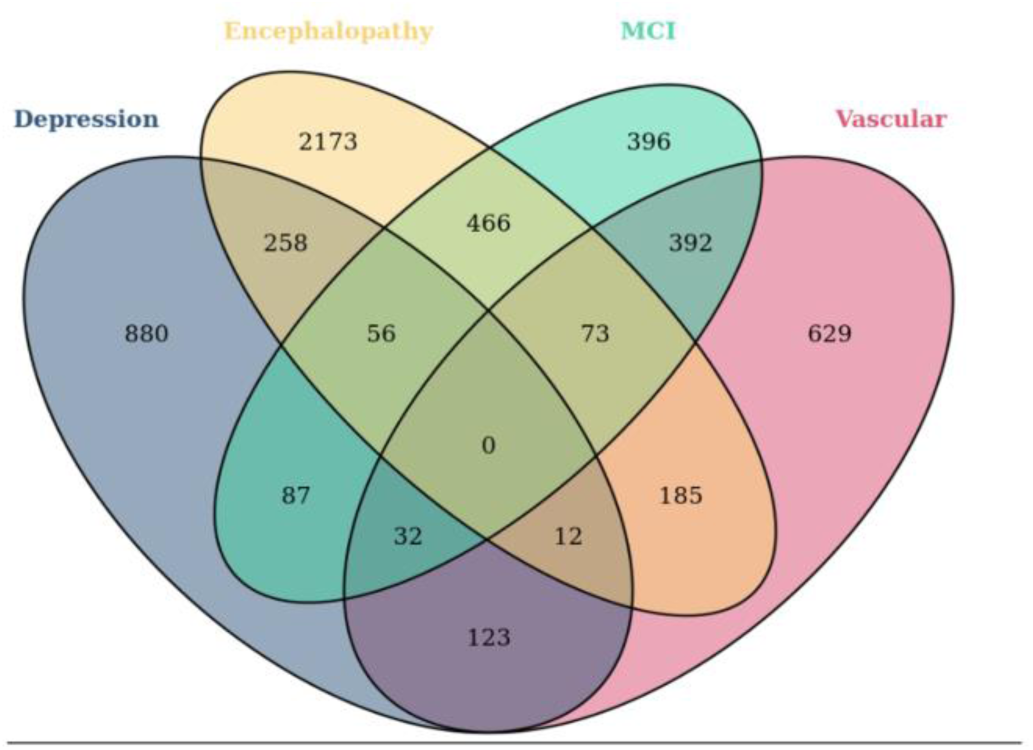
Venn diagram of AD patients in trajectory clusters.

In the 4,078 patients with unique trajectory clusters, we observed significant demographic and EHR feature differences across the four identified clusters (**Table 3**). Patients in the encephalopathy cluster were younger at their most recent visit. The depression cluster had a higher proportion of female and Hispanic patients, while the vascular disease cluster had a higher proportion of Asian patients. The depression cluster also had a higher prevalence of death. Patients in the vascular cluster generally had longer EHR records and more comorbidities. Sensitivity analyses, including all patients (multi-cluster patients included, N = 5,762), showed similar results (**Supplementary Table 2**).

**Table 3.** AD patient characteristics by trajectory clusters, patients with unique trajectory cluster only (N = 4,078)

|  | Depression | Encephalopathy | MCI | Vascular | p-value |
| --- | --- | --- | --- | --- | --- |
| **Demographics** |  |  |  |  |  |
| N with unique cluster (% total) | 880 (60.8%) | 2173 (67.4%) | 396 (26.4%) | 629 (43.5%) | <0.001 |
| Age at last visit | 82 (7) | 81.3 (7.1) | 83 (6.8) | 83.2 (7.5) | <0.001 |
| Gender (female) | 632 (71.8%) | 1,305 (60.1%) | 241 (60.9%) | 369 (58.7%) | <0.001 |
| Race-ethnicity |  |  |  |  | <0.001 |
| Amerian-Indian | 3 (0.3%) | 4 (0.2%) | 2 (0.5%) | 0 (0.0%) |  |
| Asian | 84 (9.5%) | 264 (12.1%) | 51 (12.9%) | 98 (15.6%) |  |
| Black | 43 (4.9%) | 129 (5.9%) | 24 (6.1%) | 31 (4.9%) |  |
| Hispanic | 121 (13.8%) | 219 (10.1%) | 29 (7.3%) | 63 (10.0%) |  |
| NH-White | 575 (65.3%) | 1,396 (64.2%) | 274 (69.2%) | 402 (63.9%) |  |
| Others | 54 (6.1%) | 161 (7.4%) | 16 (4.0%) | 35 (5.6%) |  |
| Death | 425 (48.3%) | 972 (44.7%) | 162 (40.9%) | 222 (35.3%) | <0.001 |
| **EHR features** |  |  |  |  |  |
| EHR record length | 5.7 (6.3) | 2.7 (4.7) | 6 (5.7) | 8.4 (4.7) | <0.001 |
| Total number of encounters | 42 (77) | 12 (26) | 36.5 (58.2) | 72 (68) | <0.001 |
| EHR record density (per year) | 10.4 (11.4) | 7.4 (11.4) | 8.3 (9.4) | 9.8 (8.8) | <0.001 |
| Total number of diagnoses | 47 (53.2) | 21 (27) | 39 (46) | 65 (38) | <0.001 |
| Location sources |  |  |  |  | <0.001 |
| UC Site 1 | 140 (15.9%) | 213 (9.8%) | 36 (9.1%) | 89 (14.1%) |  |
| UC Site 2 | 105 (11.9%) | 528 (24.3%) | 60 (15.2%) | 87 (13.8%) |  |
| UC Site 3 | 297 (33.8%) | 699 (32.2%) | 151 (38.1%) | 232 (36.9%) |  |
| UC Site 4 | 1 (0.1%) | 11 (0.5%) | 0 (0.0%) | 0 (0.0%) |  |
| UC Site 5 | 223 (25.3%) | 343 (15.8%) | 74 (18.7%) | 130 (20.7%) |  |
| UC Site 6 | 114 (13.0%) | 379 (17.5%) | 75 (18.9%) | 91 (14.5%) |  |
| **AD features** |  |  |  |  |  |
| Age at AD diagnosis | 79.3 (5.7) | 79.3 (7.1) | 80.3 (6.3) | 81 (6.9) | <0.001 |
| AD to last visit | 1.8 (3.4) | 1.2 (2.9) | 1.6 (3.1) | 1.5 (2.8) | <0.001 |

Regarding the distribution of cumulative symptoms, patients in the vascular cluster were more likely to experience symptoms involving cognition, perception, emotional state, and behavior than other clusters. However, when comparing symptoms within three years and one year before AD diagnosis, only differences in other symptoms and signs involving cognitive functions and awareness (R41), which includes memory loss (R41.3), and dizziness and giddiness (R42) were significant, with patients in the MCI and vascular clusters experiencing these symptoms more frequently (**Supplementary Table 3A**). Additionally, comparing symptoms across all systems (**Supplementary Table 3B**) revealed significant differences, with patients in the vascular disease cluster more likely to have symptoms from other systems.

In examining the features of AD, we noted significant variations in disease progression and prognosis among different clusters. Specifically, patients categorized in the vascular and MCI clusters tend to be older at the time of their initial AD diagnosis compared to those in other clusters. Those in the encephalopathy cluster, on the other hand, experienced a notably shorter duration from their first AD diagnosis to their last recorded visit, as detailed in **Table 3**.

Furthermore, survival analysis using Kaplan-Meier curves and log-rank tests revealed substantial differences in the time from first visit to AD diagnosis among the four clusters. The sequence from shortest to longest time to diagnosis was as follows: encephalopathy, depression, MCI, and vascular disease clusters. Additionally, we observed a significantly shorter duration from AD diagnosis to death in the encephalopathy cluster compared to the vascular disease cluster among deceased patients, as illustrated in **Figure 4**. Additionally, the duration from central nodes to the AD diagnosis is significantly shorter for patients in the encephalopathy cluster (G93) compared to those in other clusters (**Figure 5**).

**Figure 4.**
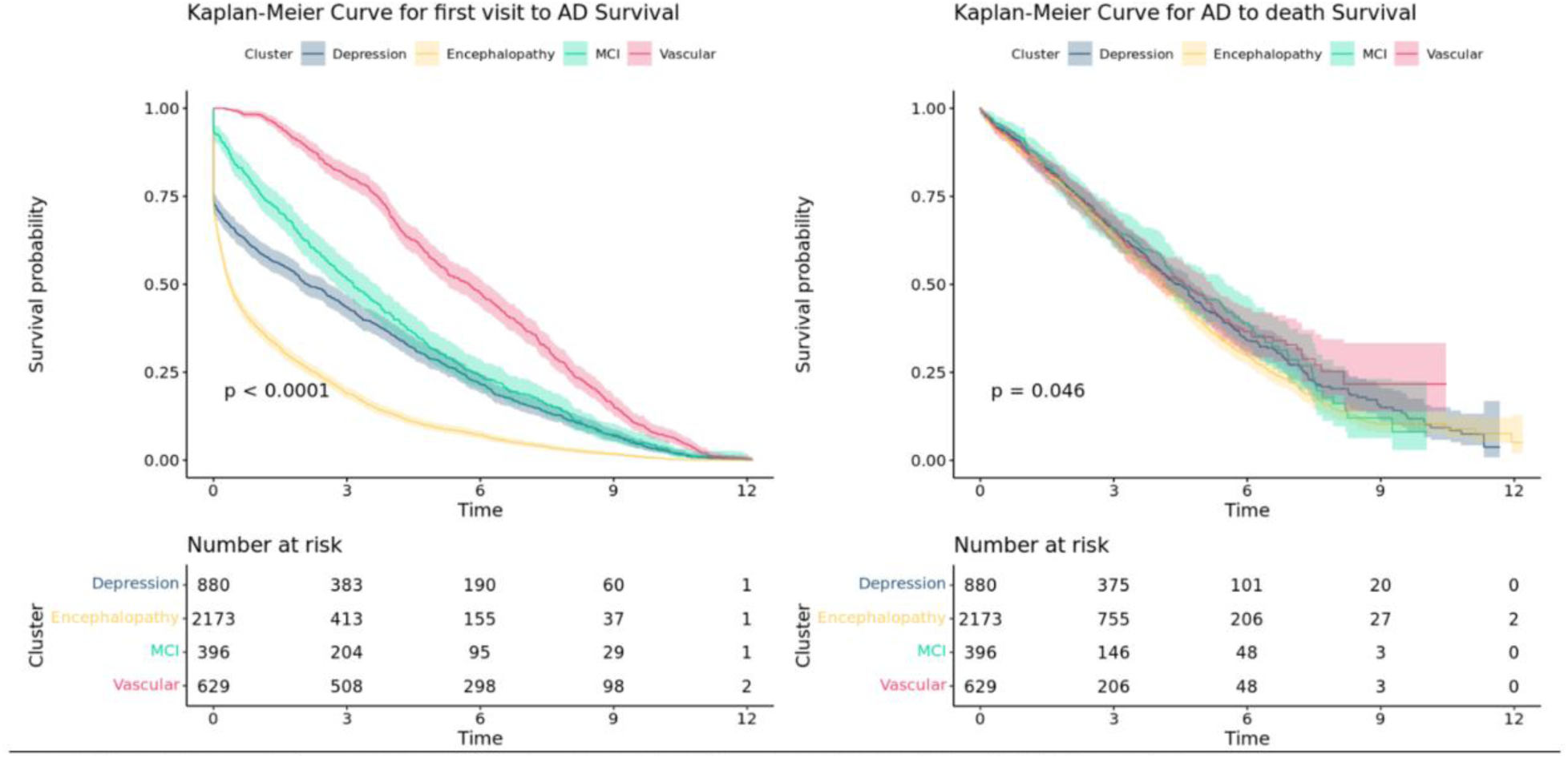
Kaplan-Meler suvival curve in AD patients by trajectory cluster. A) From first visit to AD; B) From first AD diagnosis to death.

**Figure 5.**
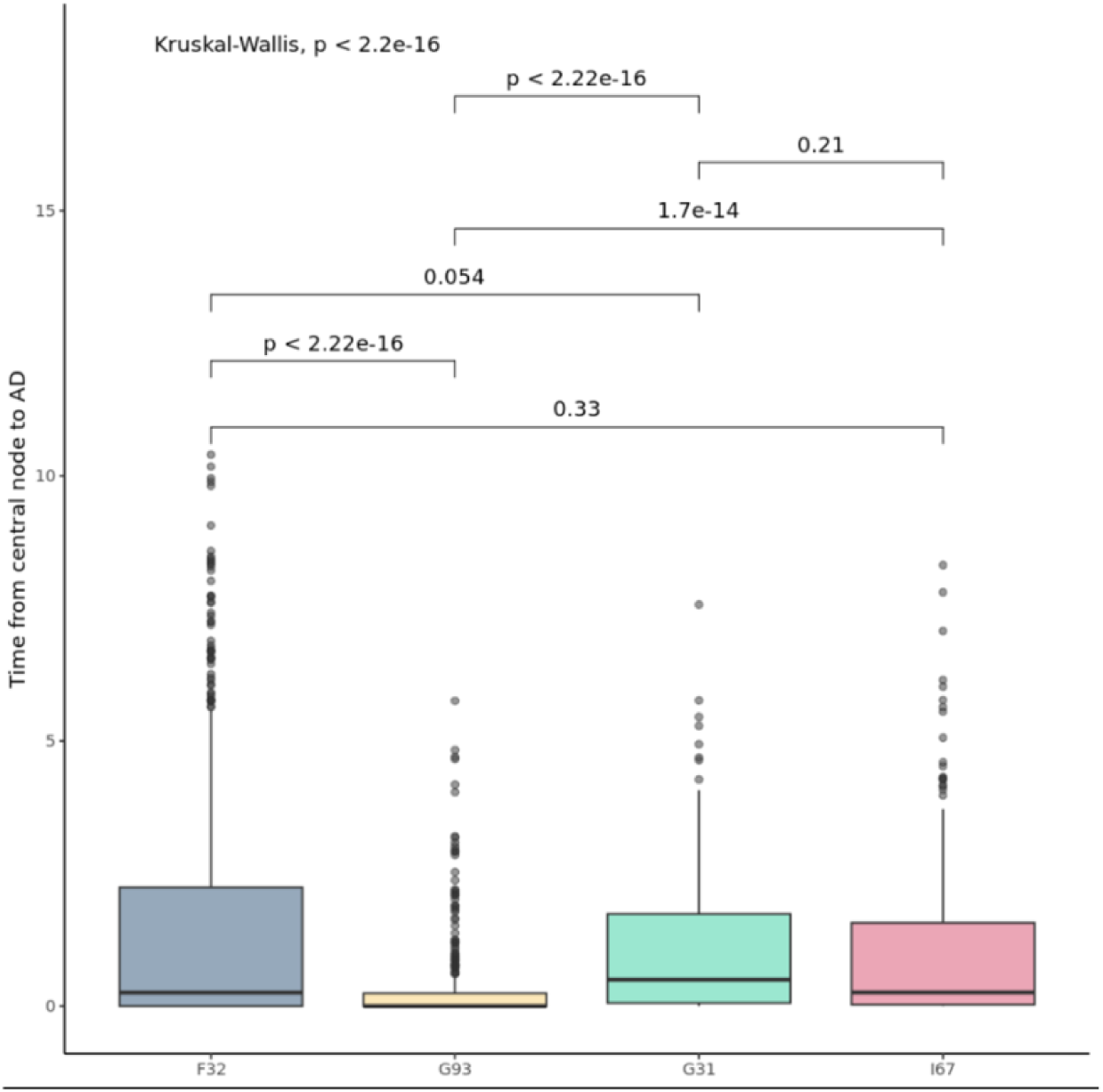
Distributions of time (in years) from central nodes to first AD diagnosis by trajectory cluster.

We further investigated potential risk factors for mortality by comparing the diagnosis distributions between alive and deceased patients within each cluster, as shown in **Supplementary Table 4**. Our focus was particularly on diagnoses that appeared more frequently among deceased patients. For instance, in the encephalopathy cluster, AD patients diagnosed with other hypothyroidism (E03), essential hypertension (I10), and other disorders of the urinary system (N39) had a higher likelihood of mortality. Similarly, in the vascular disease cluster, patients diagnosed with atrial fibrillation and flutter (I48), unspecified pneumonia (J18), and acute kidney failure (N17) also showed a higher mortality rate. These findings suggest that such diagnoses may serve as cluster-specific indicators of increased mortality risk in AD patients.

#### Single-cluster vs. multi-cluster patients

Given the findings outlined above, the MCI cluster displayed the highest proportion of patients with trajectories across multiple clusters, as depicted in **Figure 3**. Therefore, we selected the MCI cluster to compare characteristics between patients belonging to a single cluster and those appearing in multiple clusters. Patients with only MCI cluster trajectories served as the reference group. We compared these patients to those with trajectories spanning two clusters (including MCI) and three clusters (including MCI), with the results presented in **Table 4**.

**Table 4.**
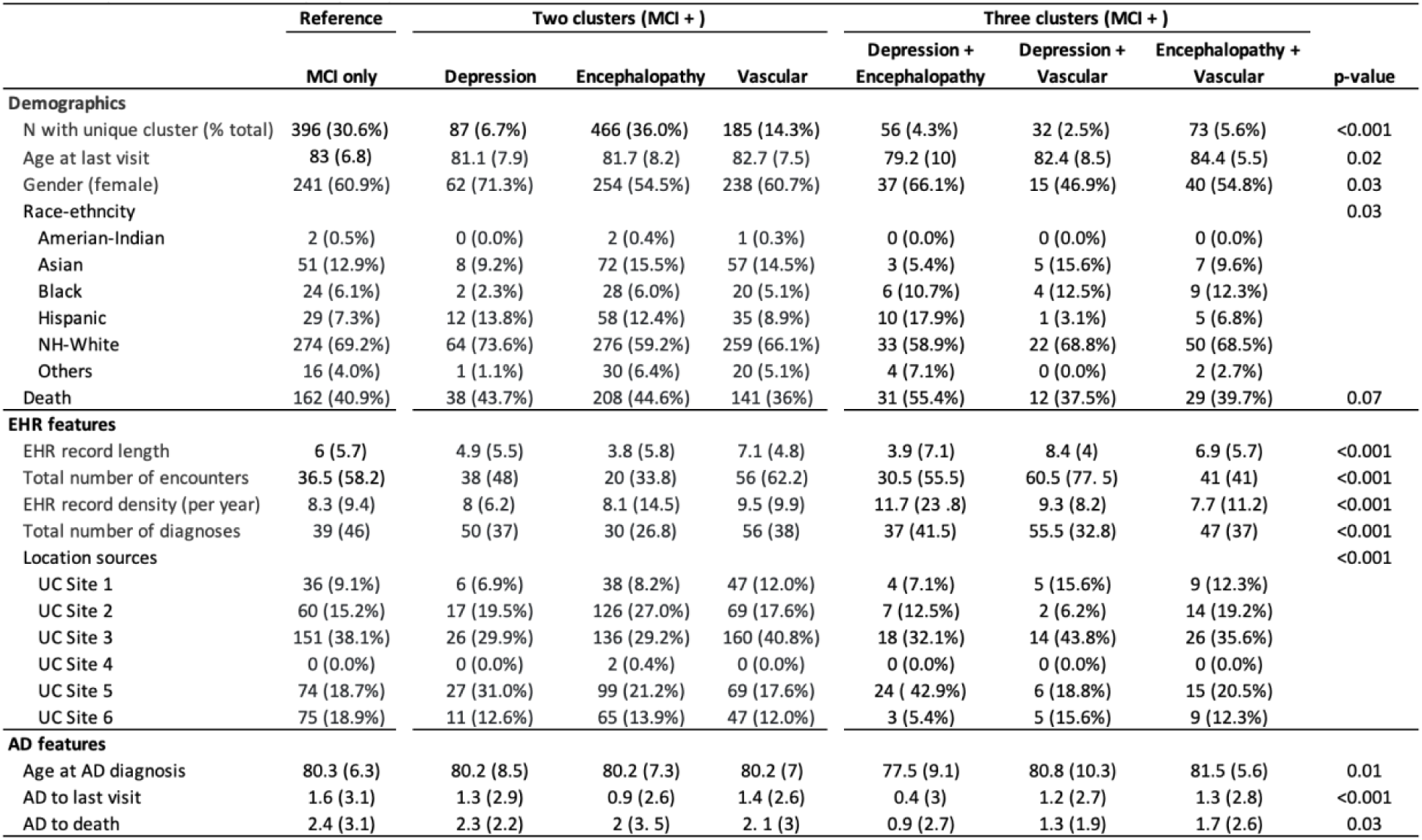
AD patient characteristics by trajectory clusters.

Notably, more than a third of the patients in the MCI cluster also had trajectories in the encephalopathy cluster. Compared to patients with only MCI trajectories, those with trajectories in multiple clusters were younger and exhibited a higher mortality rate. Consistent with single cluster comparisons shown in **Table 3**, females and individuals from the Hispanic group were more likely to have additional trajectories involving depression (e.g., two cluster depression Hispanic 13.8% vs. MCI only Hispanic 7.3%), while the Black group was more prevalent in multiple clusters compared to other racial and ethnic groups (e.g., three cluster Black 10.7-12.3% vs. MCI only Black 6.1%). Regarding AD features, patients with multi-cluster trajectories were diagnosed at a younger age and experienced a shorter duration from AD diagnosis to death (e.g., three cluster 0.9-1.7 years vs. MCI only 2.4 years).

#### Causal structural learning and risk trajectories test in controls

We employed causal structural learning with the GES algorithm to ascertain the causality of identified trajectories within the simplified networks of each cluster. Within these networks, 26.2% of the edges were deemed causal. When analyzed by cluster, the percentages were as follows: 26.3% in the depression cluster, 42.9% in the encephalopathy cluster, 28.2% in the mild MCI cluster, and 24.1% in the vascular cluster. Details of these causal links are provided in **Supplementary Figure 4**. Among the AD risk factors identified through the FG model, six ICD codes were potentially causally associated with AD. These include unspecified dementia (F03), vascular dementia (F01), depressive episode (F32), unspecified mood disorder (F39), other disorders of the brain (G93), and other cerebrovascular diseases (I67). Notably, three of these codes (F32, G93, and I67) were identified as central nodes within each respective cluster.

Finally, we applied a similar FG proportional subdistribution hazards model framework to test the top common trajectories of each cluster in a case-control sample, as detailed in **Table 5**. Our initial analysis focused on the associations between identified trajectories and AD, aiming to validate our hypothesis that these trajectories represent true risk factors for AD. We found that all representative risk trajectories in each cluster were positively associated with AD; in other words, patients with a given trajectory exhibited higher risks of developing AD compared to those without this trajectory. Subsequently, we compared the effect sizes between multi-step trajectories and single risk factors by evaluating the differences between patients with the risk trajectory and those possessing any of the factors within the trajectory. The results indicated that patients with most of these trajectories faced higher risks of AD compared to those with only a single risk factor.

**Table 5.** Associations between risk trajectories and incidence of Alzheimer’s disease.

| Cluster | Risk trajectory | Risk trajectory vs. all others |  |  | Risk trajectory vs. single factor |  |  |
| --- | --- | --- | --- | --- | --- | --- | --- |
|  |  | Sample N | RR | P-value | Sample N | RR | P-value |
| Depression<br>(Central node - F32) | E78 → F32 | 43889 | 1.40 | <0.001 | 24611 | 1.61 | <0.001 |
|  | F41 → F32 | 43889 | 1.57 | <0.001 | 8970 | 1.10 | 0.01 |
|  | I10 → F32 | 43889 | 1.47 | <0.001 | 26257 | 1.67 | <0.001 |
|  | E11 → F32 | 43889 | 1.54 | <0.001 | 14502 | 1.39 | <0.001 |
|  | M54 → F32 | 43889 | 1.34 | <0.001 | 17015 | 1.50 | <0.001 |
|  | K59 → F32 | 43889 | 1.49 | <0.001 | 11625 | 1.24 | <0.001 |
| Encephalopathy<br>(Central node - G93) | E78 → G93 | 43889 | 1.66 | <0.001 | 25288 | 1.87 | <0.001 |
|  | I10 → G93 | 43889 | 1.66 | <0.001 | 26836 | 1.82 | <0.001 |
|  | E11 → G93 | 43889 | 1.74 | <0.001 | 12029 | 1.71 | <0.001 |
|  | N39 → G93 | 43889 | 1.69 | <0.001 | 11552 | 1.61 | <0.001 |
|  | N40 → G93 | 43889 | 1.66 | <0.001 | 8957 | 1.81 | <0.001 |
|  | N17 → G93 | 43889 | 1.67 | <0.001 | 6181 | 1.33 | <0.001 |
| MCI<br>(Central node - G31) | G45 → G31* | 43889 | 2.30 | <0.001 | 6837 | 0.87 | 0.002 |
|  | N95 → G31* | 43889 | 2.39 | <0.001 | 8587 | 1.60 | <0.001 |
|  | N52 → G31* | 43889 | 2.62 | <0.001 | 7356 | 1.47 | <0.001 |
|  | H01 → G31* | 43889 | 2.26 | <0.001 | 8079 | 1.15 | <0.001 |
|  | H04 → G31* | 43889 | 2.20 | <0.001 | 10422 | 1.60 | <0.001 |
| Vascular<br>(Central node - I67) | E78 → I67 | 43889 | 1.62 | <0.001 | 25120 | 1.82 | <0.001 |
|  | I10 → I67 | 43889 | 1.67 | <0.001 | 26763 | 1.80 | <0.001 |
|  | M25 → I67 | 43889 | 1.58 | <0.001 | 17517 | 1.96 | <0.001 |
|  | G31 → I67* | 43889 | 2.82 | <0.001 | 6699 | 1.08 | 0.04 |
|  | Z98 → I67 | 43889 | 1.46 | <0.001 | 13483 | 2.06 | <0.001 |
|  | M79 → I67 | 43889 | 1.57 | <0.001 | 15296 | 1.98 | <0.001 |
|  | D64 → I67 | 43889 | 1.47 | <0.001 | 10847 | 1.46 | <0.001 |

**Notes:**
\* Model experienced complete separation, where the AD cases separates the predictor variable completely. Parameter estimates and p-values might be incorrect.

## Discussion

In our study, we analyzed common disease patterns of AD using DTW, clustering, and network analyses based on EHRs from the University of California Health Data Warehouse. We used longitudinal records from 5,762 eligible AD patients to identify four distinct clusters of disease trajectories: the mental health cluster, the encephalopathy cluster, the MCI cluster, and the vascular disease cluster. Each cluster exhibited unique characteristics, along with some shared diagnoses and pathways. We rigorously evaluated these trajectories through association tests, comparison to controls, and causal structural learning, contributing new insights to the existing literature on AD. This comprehensive approach offers new insights into the temporal dynamics of AD progression and highlights critical windows for targeted interventions that could alter the disease’s trajectory.

The four clusters of AD trajectories identified in our study align with prior research linking mental illness, brain diseases, and vascular diseases to AD risk. Previous studies have demonstrated that conditions like depression are linked to an increased risk of AD [5,40], as they may induce neurodegenerative changes predisposing individuals to the disease. For instance, mechanisms related to major depression, such as chronic inflammation and the hyperactivation of the hypothalamic–pituitary–adrenal axis, have been implicated in the pathogenesis of AD [41]. Similarly, encephalopathy is associated with progressive neurodegeneration and increased AD risk due to pathological changes like hyperphosphorylated tau [42]. Moreover, vascular conditions such as hypertension and cardiovascular diseases contribute to compromised cerebral blood flow and metabolic dysfunction, heightening AD risk [43].

However, these earlier studies typically focused on individual risk factors and their direct associations with AD, often overlooking the sequential and cumulative impact of these conditions. Our research builds on this foundation by examining the complete spectrum of diseases potentially leading to AD, marking a novel approach in systematically exploring the multi-step disease progression. We are the first to incorporate the sequence of disease onset into AD research, underscoring that the timing and order of risk factors are crucial. Our findings suggest that understanding and predicting disease progression could significantly enhance intervention strategies. For example, we identified a possible trajectory of acute kidney failure (N17) → unspecified encephalopathy (G93) → AD (G30). Thus, monitoring kidney function and managing related symptoms could potentially delay or prevent the onset of neurological symptoms. Likewise, early mental health interventions might be prioritized if depression is identified before the onset of significant memory loss, potentially delaying or reducing the severity of AD. In addition, while it is well-established that cardiovascular disease can precede cognitive impairments [43], trajectories we identified from the vascular cluster (e.g. unspecified encephalopathy (G93) → other cerebrovascular disease (I67) → AD (G30)) suggest that specific sequences of vascular conditions might sharpen the focus on particular patient subgroups who are at an elevated risk for AD. For instance, the presence of cerebrovascular disease alone signals a risk, necessitating interventions such as blood pressure management. However, if a patient also has a history of encephalopathy prior to cerebrovascular issues, this might warrant an even more rigorous approach to managing these risk factors. Recognizing such detailed pathways not only enhances our understanding of disease progression but also facilitates personalized medical strategies. This could lead to earlier and more aggressive management of cardiovascular health in those at heightened risk for AD, ultimately improving outcomes.

Our study adopts a unique approach to preprocessing patient records before extracting AD trajectories, distinguishing it from much of the existing trajectory research. EHRs contain a patient’s comprehensive health history, which over the years, can accumulate into thousands of entries. However, many of these entries may not relate to AD and could introduce noise, complicating the analysis—especially given the smaller size of our AD dataset [44]. Among the AD patients we studied (N = 24,473), there was considerable variability in the number of unique three-digit ICD codes, ranging from one to 90, with a median of 40.5. This variability makes it challenging to compare patient trajectories directly. To address this, we implemented a cleaning process to simplify these trajectories by focusing only on positively associated ICD pairs identified from the FG models, thus retaining only those diagnostic patterns considered risky.

Additionally, we sampled trajectories for patients with an extensive number of trajectories to minimize bias towards those with longer or more detailed histories. This preprocessing ensures that we maintain crucial diagnostic patterns, enhancing the quality and interpretability of the trajectories we analyze. This methodological rigor is essential for accurately interpreting the complex pathways leading to AD.

We introduced another innovative aspect by applying clustering methods to AD trajectories. Clustering is crucial because it reveals significant differences in demographics, accompanying symptoms, and the progression of AD among the groups. Recognizing these distinctions is essential for understanding the multifaceted nature of AD, enabling more targeted research, and potentially informing more personalized treatment approaches. For example, differences in gender and race-ethnicity distributions across different clusters can lead to tailored public health initiatives and interventions that address specific risk factors and disease manifestations in diverse populations. Moreover, significant variations in AD progression and prognosis among different clusters, such as faster time to death in the encephalopathy cluster, highlight the need for differential treatment strategies and care plans that are sensitive to the expected disease trajectory. On the other hand, the fact that patients categorized in the MCI clusters tend to be older at the time of their initial AD diagnosis compared to those in other clusters suggests the possibility of late-onset disease management strategies and preventive measures that could delay the onset or progression of AD in at-risk older adults.

Finally, to assess the robustness of our results, we rigorously evaluated the identified AD trajectories using association tests and causal structural learning. Association tests confirmed that patients with identified AD risk trajectories had higher risks of developing AD compared to those without these trajectories. Importantly, these multi-step trajectories posed greater risks than individual risk factors. This underscores the significance of examining disease risk factors collectively and sequentially. However, the results from causal structural learning are more exploratory in nature.

Our study faces limitations. Unlike survey studies where data collection follows a regular schedule, patients do not visit hospitals consistently, which challenges the chronological capture of disease progression in EHRs. Multiple diagnoses are often recorded in a single visit, which may not accurately reflect the intrinsic order of disease progression. Another limitation is that the different clustering methods we used showed moderate agreement (adjusted rand indices: 0.13-0.23), which is not surprising given the complexity of the data and the fact that many individuals fell into multiple clusters—1,511 in two clusters and 173 in three clusters. This variability in clustering outcomes suggests that there may not be distinct boundaries between different clusters, reflecting the likelihood that individuals may experience multiple comorbidities before the onset of AD. However, the absence of clear cluster boundaries does not diminish the importance of identifying these clusters. There are also limitations to causal structure learning and related methods to identify true causal relationship as they performed in observational data.

In conclusion, our study has successfully identified and analyzed distinct progression patterns in AD by utilizing a comprehensive computational framework to examine longitudinal trajectories. This methodological advancement enables us to delve beyond the traditional analysis of isolated comorbidities, providing a deeper understanding of the complex interrelations and temporal dynamics that characterize AD progression. The insights gained from our detailed trajectory analysis enhance our understanding of AD, potentially improving diagnostic accuracy and enabling the development of more effective preventative strategies. By elucidating the pathways of AD progression and their causal relationships, our research lays a solid foundation for future studies aimed at designing targeted interventions. These could significantly enhance patient care and outcomes by addressing the disease’s complexity and tailoring medical and care strategies to meet the specific needs associated with different AD progression types. This innovative approach opens new avenues for research and offers significant potential for advancing patient care in AD.

## Data Availability

All data produced in the present study are available upon reasonable request to the authors.

## Acknowledgements

We thank L. Dahm and the Center for Data-driven Insights and Innovation at UC Health (CDI2; www.ucop.edu/uc-health/functions/center-for-data-driven-insights-and-innovationscdi2.html) for analytical and technical support related to use of the UC Health Data Warehouse.

---

**Supplementary Figure 1.**
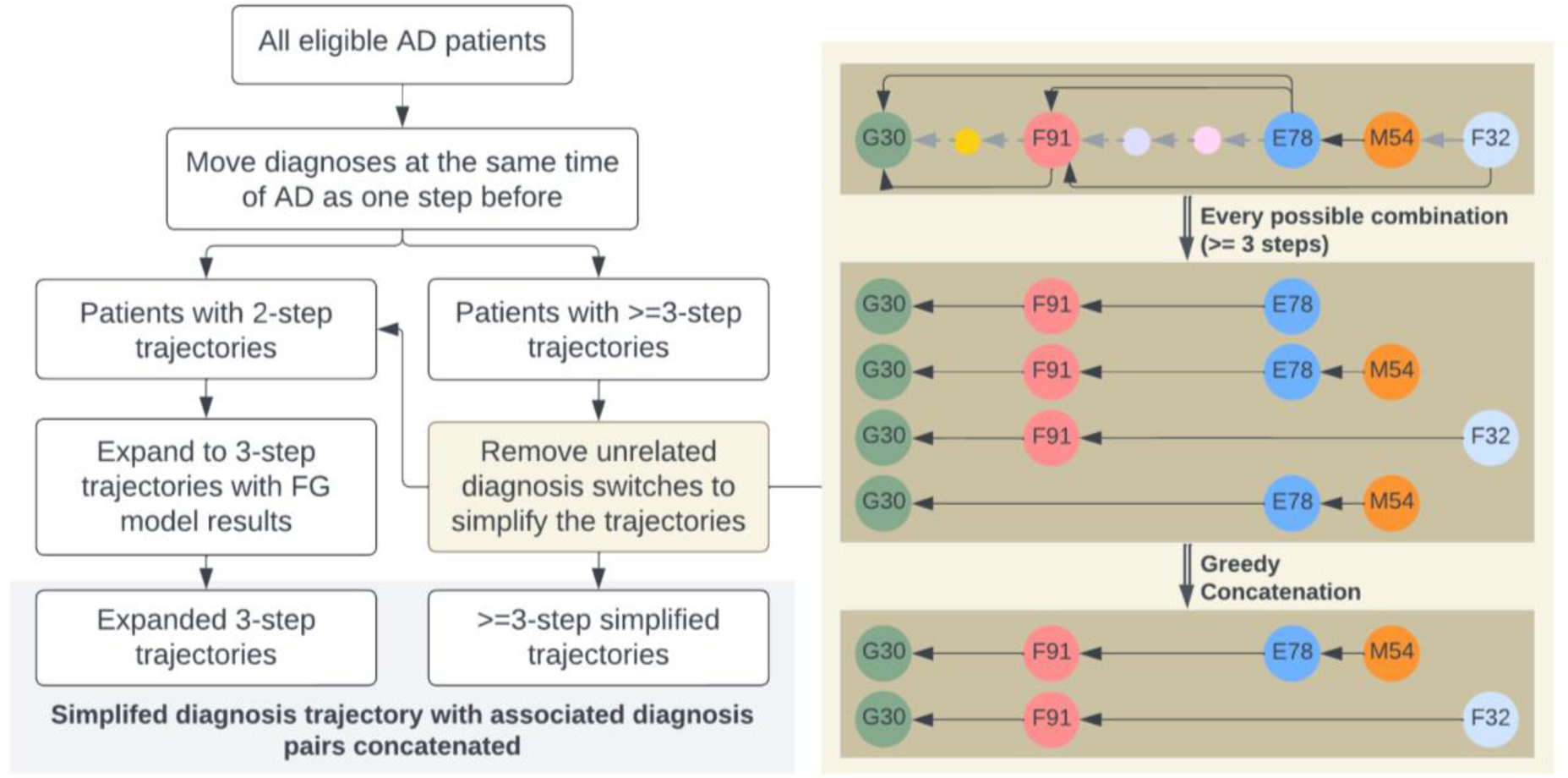
Simplify trajectories.

**Supplementary Figure 2.**
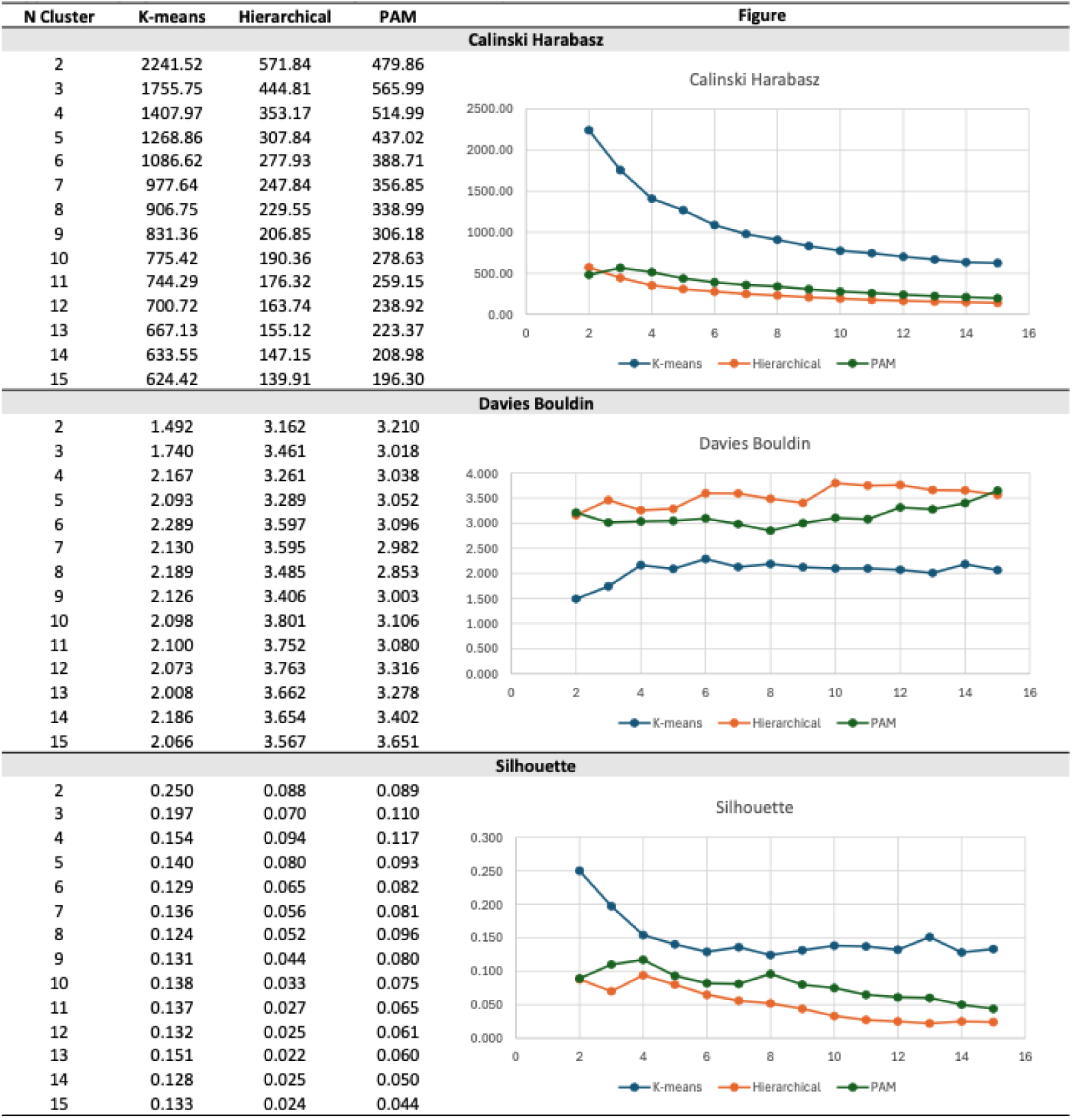
Determine clustering method and the optimal number of clusters

**Supplementary Figure 3.**
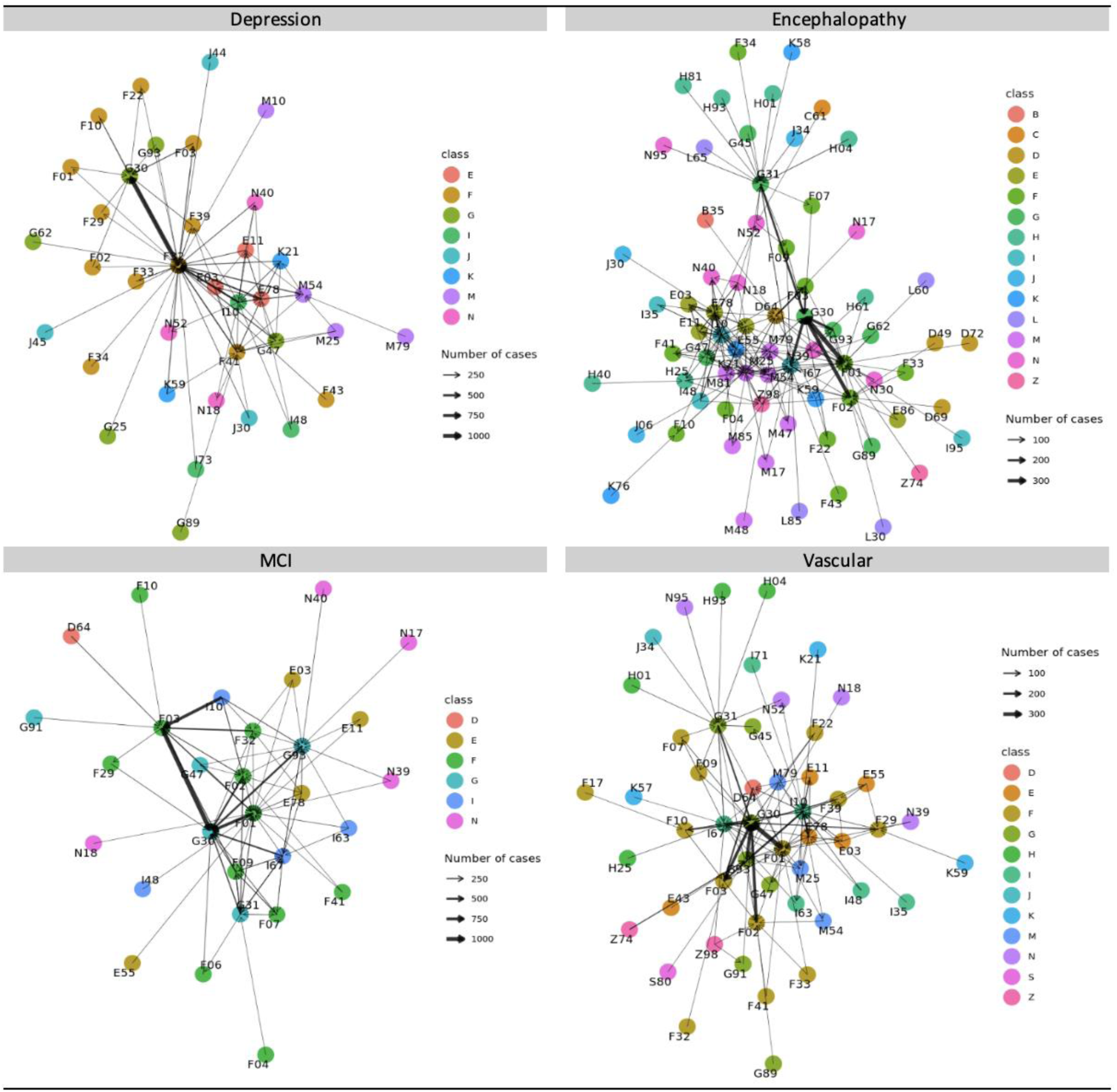
Raw networks by AD trajectory clusters. Shows connections followed by at least 0.5% of patient in each cluster.

**Supplementary Table 1.** Descriptive statistics and matching information of AD cases vs. 1:1 propensity score-matched non-AD controls in UC data warehouse (N = 48,946)

| Characteristic | AD<br>N = 24,473 | Non-AD<br>N = 24,473 | Mean Diff.<br>before | Mean Diff.<br>after |
| --- | --- | --- | --- | --- |
| Age at last encounter visit | 81.12 (5.4) | 81.3 (5.6) | 1.0091 | -0.0246 |
| Gender (female) | 15164 (62.0%) | 15018 (61.4%) | 0.1367 | 0.0123 |
| Race-ethnicity |  |  |  |  |
| Amerian-Indian | 62 (0.3%) | 51 (0.2%) | -0.0050 | 0.0089 |
| Asian | 2940 (12.0%) | 2599 (10.6%) | 0.0141 | 0.0429 |
| Black | 1353 (5.5%) | 1139 (4.7%) | 0.0374 | 0.0383 |
| Hispanic | 2775 (11.3%) | 2507 (10.2%) | 0.0451 | 0.0345 |
| NH-White | 15891 (64.9%) | 16647 (68.0%) | -0.0879 | -0.0647 |
| Others | 1452 (5.9%) | 1530 (6.3%) | 0.0624 | -0.0135 |
| Location resource |  |  |  |  |
| UC Site 1 | 3465 (14.2%) | 3449 (14.1%) | -0.0692 | 0.0019 |
| UC Site 2 | 3575 (14.6%) | 3307 (13.5%) | 0.1418 | 0.0310 |
| UC Site 3 | 8123 (33.2%) | 8602 (35.2%) | -0.0423 | -0.0416 |
| UC Site 4 | 71 (0.3%) | 47 (0.2%) | 0.0296 | 0.0182 |
| UC Site 5 | 5168 (21.1%) | 4815 (19.7%) | 0.0416 | 0.0353 |
| UC Site 6 | 4071 (16.6%) | 4253 (17.4%) | -0.0662 | -0.0200 |

**Supplementary Table 2.**
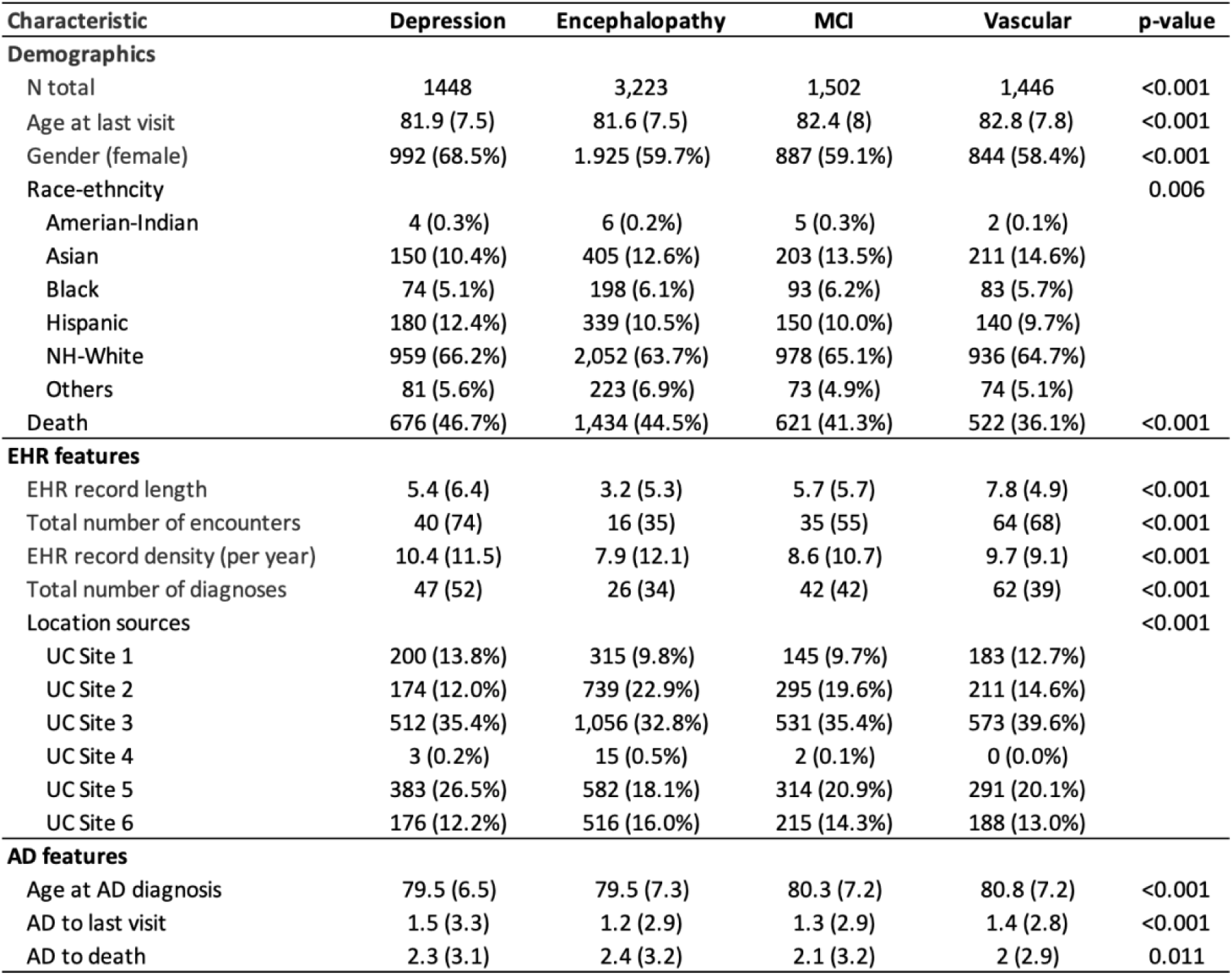
AD patient characteristics by trajectory clusters, full patients in each cluster (N = 5,762)

| Characteristic | Depression | Encephalopathy | MCI | Vascular | p-value |
| --- | --- | --- | --- | --- | --- |
| **Demographics** |  |  |  |  |  |
| N total | 1448 | 3,223 | 1,502 | 1,446 | <0.001 |
| Age at last visit | 81.9 (7.5) | 81.6 (7.5) | 82.4 (8) | 82.8 (7.8) | <0.001 |
| Gender (female) | 992 (68.5%) | 1,925 (59.7%) | 887 (59.1%) | 844 (58.4%) | <0.001 |
| Race-ethnicity |  |  |  |  | 0.006 |
| Amerian-Indian | 4 (0.3%) | 6 (0.2%) | 5 (0.3%) | 2 (0.1%) |  |
| Asian | 150 (10.4%) | 405 (12.6%) | 203 (13.5%) | 211 (14.6%) |  |
| Black | 74 (5.1%) | 198 (6.1%) | 93 (6.2%) | 83 (5.7%) |  |
| Hispanic | 180 (12.4%) | 339 (10.5%) | 150 (10.0%) | 140 (9.7%) |  |
| NH-White | 959 (66.2%) | 2,052 (63.7%) | 978 (65.1%) | 936 (64.7%) |  |
| Others | 81 (5.6%) | 223 (6.9%) | 73 (4.9%) | 74 (5.1%) |  |
| Death | 676 (46.7%) | 1,434 (44.5%) | 621 (41.3%) | 522 (36.1%) | <0.001 |
| **EHR features** |  |  |  |  |  |
| EHR record length | 5.4 (6.4) | 3.2 (5.3) | 5.7 (5.7) | 7.8 (4.9) | <0.001 |
| Total number of encounters | 40 (74) | 16 (35) | 35 (55) | 64 (68) | <0.001 |
| EHR record density (per year) | 10.4 (11.5) | 7.9 (12.1) | 8.6 (10.7) | 9.7 (9.1) | <0.001 |
| Total number of diagnoses | 47 (52) | 26 (34) | 42 (42) | 62 (39) | <0.001 |
| Location sources |  |  |  |  | <0.001 |
| UC Site 1 | 200 (13.8%) | 315 (9.8%) | 145 (9.7%) | 183 (12.7%) |  |
| UC Site 2 | 174 (12.0%) | 739 (22.9%) | 295 (19.6%) | 211 (14.6%) |  |
| UC Site 3 | 512 (35.4%) | 1,056 (32.8%) | 531 (35.4%) | 573 (39.6%) |  |
| UC Site 4 | 3 (0.2%) | 15 (0.5%) | 2 (0.1%) | 0 (0.0%) |  |
| UC Site 5 | 383 (26.5%) | 582 (18.1%) | 314 (20.9%) | 291 (20.1%) |  |
| UC Site 6 | 176 (12.2%) | 516 (16.0%) | 215 (14.3%) | 188 (13.0%) |  |
| **AD features** |  |  |  |  |  |
| Age at AD diagnosis | 79.5 (6.5) | 79.5 (7.3) | 80.3 (7.2) | 80.8 (7.2) | <0.001 |
| AD to last visit | 1.5 (3.3) | 1.2 (2.9) | 1.3 (2.9) | 1.4 (2.8) | <0.001 |
| AD to death | 2.3 (3.1) | 2.4 (3.2) | 2.1 (3.2) | 2 (2.9) | 0.011 |

**Supplementary Table 3A.**
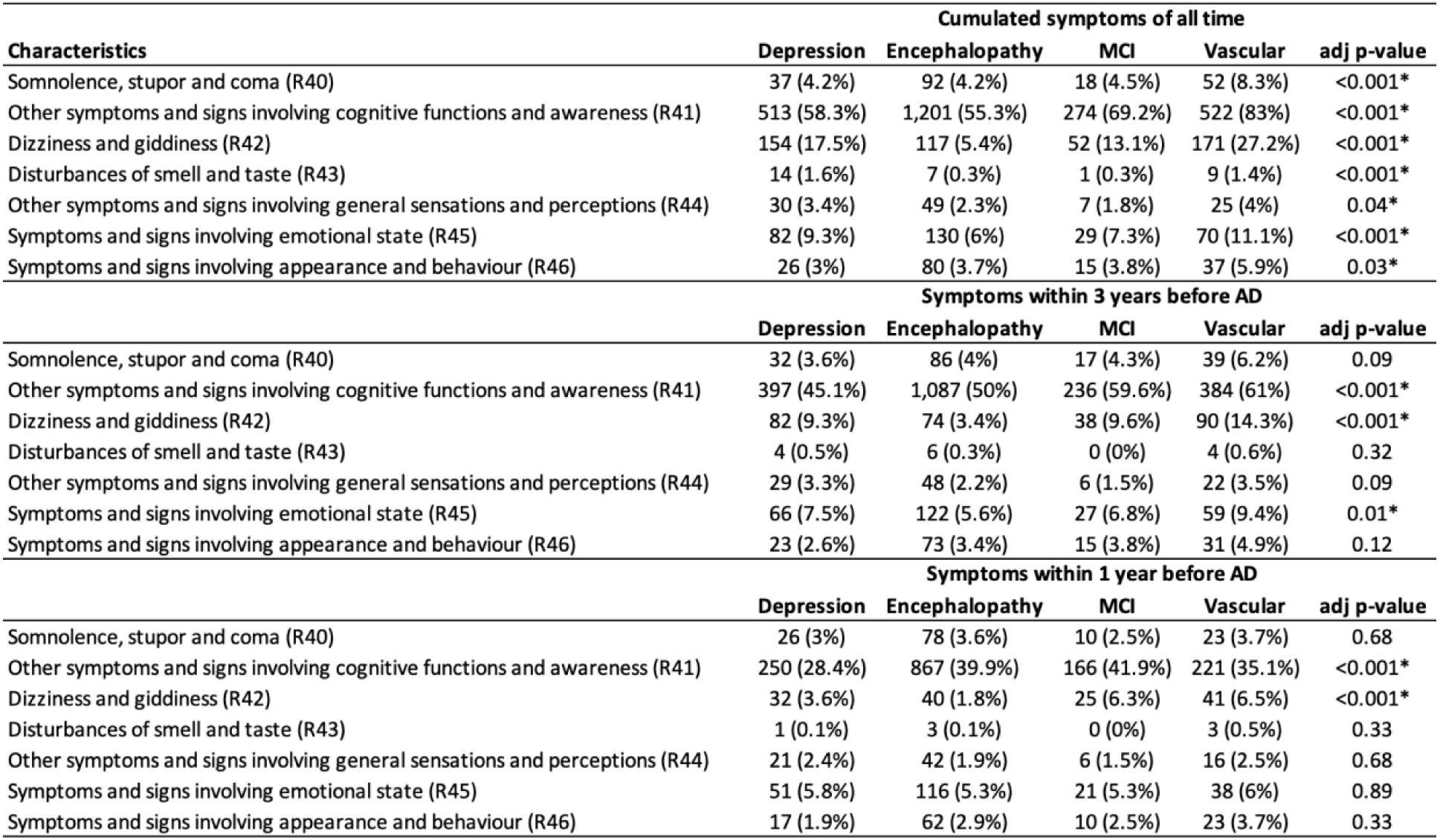
Patient cumulated symptoms involving cognition, perception, emotional state and behaviour (selected significant different in distributions) before AD diagnosis by trajectory clusters.

| Characteristics | Cumulated symptoms of all time |  |  |  |  |
| --- | --- | --- | --- | --- | --- |
|  | Depression | Encephalopathy | MCI | Vascular | adj p-value |
| Somnolence, stupor and coma (R40) | 37 (4.2%) | 92 (4.2%) | 18 (4.5%) | 52 (8.3%) | <0.001* |
| Other symptoms and signs involving cognitive functions and awareness (R41) | 513 (58.3%) | 1,201 (55.3%) | 274 (69.2%) | 522 (83%) | <0.001* |
| Dizziness and giddiness (R42) | 154 (17.5%) | 117 (5.4%) | 52 (13.1%) | 171 (27.2%) | <0.001* |
| Disturbances of smell and taste (R43) | 14 (1.6%) | 7 (0.3%) | 1 (0.3%) | 9 (1.4%) | <0.001* |
| Other symptoms and signs involving general sensations and perceptions (R44) | 30 (3.4%) | 49 (2.3%) | 7 (1.8%) | 25 (4%) | 0.04* |
| Symptoms and signs involving emotional state (R45) | 82 (9.3%) | 130 (6%) | 29 (7.3%) | 70 (11.1%) | <0.001* |
| Symptoms and signs involving appearance and behaviour (R46) | 26 (3%) | 80 (3.7%) | 15 (3.8%) | 37 (5.9%) | 0.03* |
| Characteristics | Symptoms within 3 years before AD |  |  |  |  |
|  | Depression | Encephalopathy | MCI | Vascular | adj p-value |
| Somnolence, stupor and coma (R40) | 32 (3.6%) | 86 (4%) | 17 (4.3%) | 39 (6.2%) | 0.09 |
| Other symptoms and signs involving cognitive functions and awareness (R41) | 397 (45.1%) | 1,087 (50%) | 236 (59.6%) | 384 (61%) | <0.001* |
| Dizziness and giddiness (R42) | 82 (9.3%) | 74 (3.4%) | 38 (9.6%) | 90 (14.3%) | <0.001* |
| Disturbances of smell and taste (R43) | 4 (0.5%) | 6 (0.3%) | 0 (0%) | 4 (0.6%) | 0.32 |
| Other symptoms and signs involving general sensations and perceptions (R44) | 29 (3.3%) | 48 (2.2%) | 6 (1.5%) | 22 (3.5%) | 0.09 |
| Symptoms and signs involving emotional state (R45) | 66 (7.5%) | 122 (5.6%) | 27 (6.8%) | 59 (9.4%) | 0.01* |
| Symptoms and signs involving appearance and behaviour (R46) | 23 (2.6%) | 73 (3.4%) | 15 (3.8%) | 31 (4.9%) | 0.12 |
| Characteristics | Symptoms within 1 year before AD |  |  |  |  |
|  | Depression | Encephalopathy | MCI | Vascular | adj p-value |
| Somnolence, stupor and coma (R40) | 26 (3%) | 78 (3.6%) | 10 (2.5%) | 23 (3.7%) | 0.68 |
| Other symptoms and signs involving cognitive functions and awareness (R41) | 250 (28.4%) | 867 (39.9%) | 166 (41.9%) | 221 (35.1%) | <0.001* |
| Dizziness and giddiness (R42) | 32 (3.6%) | 40 (1.8%) | 25 (6.3%) | 41 (6.5%) | <0.001* |
| Disturbances of smell and taste (R43) | 1 (0.1%) | 3 (0.1%) | 0 (0%) | 3 (0.5%) | 0.33 |
| Other symptoms and signs involving general sensations and perceptions (R44) | 21 (2.4%) | 42 (1.9%) | 6 (1.5%) | 16 (2.5%) | 0.68 |
| Symptoms and signs involving emotional state (R45) | 51 (5.8%) | 116 (5.3%) | 21 (5.3%) | 38 (6%) | 0.89 |
| Symptoms and signs involving appearance and behaviour (R46) | 17 (1.9%) | 62 (2.9%) | 10 (2.5%) | 23 (3.7%) | 0.33 |

**Supplementary Table 3B.**
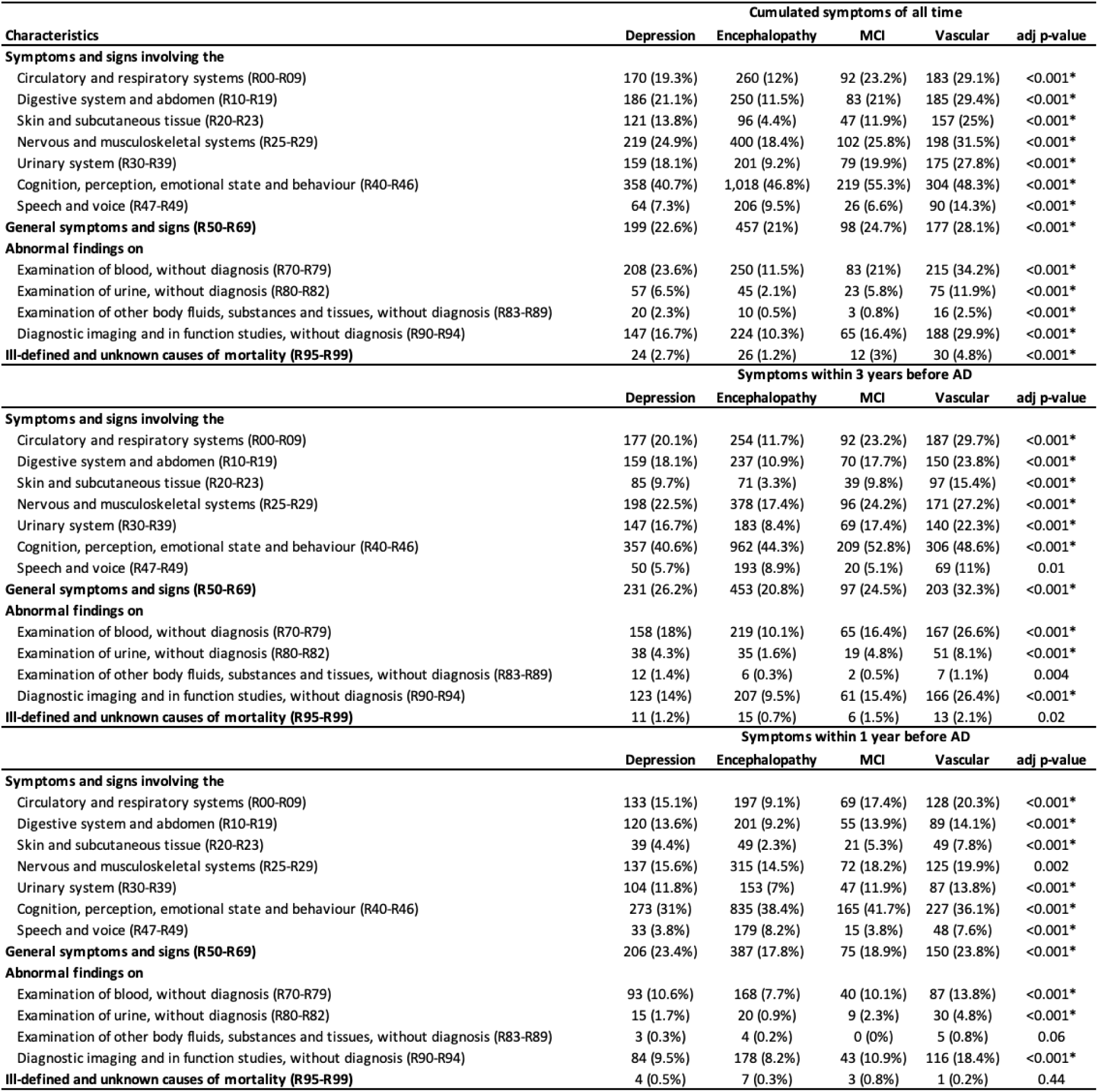
Patient cumulated symptoms (selected significant different in distributions) before AD diagnosis by trajectory dusters.

**Supplementary Table 4.** Comparisons of diagnosis in alive vs. deceased AD patients by trajectory cluster, significant diagnoses only.

| Depression (N = 880) |  |  |  | Encephalopathy (N = 2,173) |  |  |  | Vascular (N = 629) |  |  |  |
| --- | --- | --- | --- | --- | --- | --- | --- | --- | --- | --- | --- |
| ICD | Alive | Deceased | Adj_P_value | ICD | Alive | Deceased | Adj_P_value | ICD | Alive | Deceased | Adj_P_value |
| F33 | 82 (18%) | 44 (10.4%) | 0.01 | E03 | 122 (10.2%) | 134 (13.8%) | 0.04 | E11 | 130 (31.9%) | 47 (21.2%) | 0.07 |
| F41 | 195 (42.9%) | 120 (28.2%) | <0.001 | F01 | 251 (20.9%) | 245 (25.2%) | 0.05 | F01 | 85 (20.9%) | 73 (32.9%) | 0.03 |
| G31 | 142 (31.2%) | 73 (17.2%) | <0.001 | F02 | 220 (18.3%) | 72 (7.4%) | <0.001 | F02 | 110 (27%) | 27 (12.2%) | 0.002 |
| G47 | 151 (33.2%) | 106 (24.9%) | 0.05 | F03 | 674 (56.1%) | 608 (62.6%) | 0.01 | F03 | 181 (44.5%) | 128 (57.7%) | 0.04 |
| G89 | 128 (28.1%) | 67 (15.8%) | 0.001 | F09 | 236 (19.7%) | 154 (15.8%) | 0.05 | I48 | 82 (20.1%) | 66 (29.7%) | 0.09 |
| H25 | 65 (14.3%) | 37 (8.7%) | 0.06 | G31 | 510 (42.5%) | 286 (29.4%) | <0.001 | J18 | 34 (8.4%) | 37 (16.7%) | 0.04 |
| J06 | 57 (12.5%) | 31 (7.3%) | 0.06 | G47 | 171 (14.2%) | 100 (10.3%) | 0.02 | N17 | 40 (9.8%) | 42 (18.9%) | 0.04 |
| M47 | 59 (13%) | 33 (7.8%) | 0.07 | I10 | 497 (41.4%) | 486 (50%) | <0.001 | R39 | 59 (14.5%) | 16 (7.2%) | 0.098 |
| M51 | 76 (16.7%) | 35 (8.2%) | 0.02 | I67 | 249 (20.7%) | 162 (16.7%) | 0.05 | R63 | 89 (21.9%) | 72 (32.4%) | 0.07 |
| M79 | 138 (30.3%) | 91 (21.4%) | 0.02 | N39 | 112 (9.3%) | 129 (13.3%) | 0.02 | Z01 | 182 (44.7%) | 61 (27.5%) | 0.002 |
| M85 | 81 (17.8%) | 39 (9.2%) | 0.02 | R26 | 163 (13.6%) | 168 (17.3%) | 0.05 | Z11 | 85 (20.9%) | 25 (11.3%) | 0.05 |
| R10 | 96 (21.1%) | 64 (15.1%) | 0.09 | R41 | 717 (59.7%) | 484 (49.8%) | <0.001 | Z12 | 195 (47.9%) | 68 (30.6%) | 0.002 |
| R35 | 80 (17.6%) | 51 (12%) | 0.06 |  |  |  |  | Z13 | 174 (42.8%) | 59 (26.6%) | 0.003 |
| R41 | 296 (64.1%) | 217 (51.1%) | <0.001 |  |  |  |  | Z20 | 75 (18.4%) | 22 (9.9%) | 0.07 |
| R42 | 93 (20.4%) | 61 (14.4%) | 0.05 |  |  |  |  | Z66 | 29 (7.1%) | 38 (17.1%) | 0.01 |
| Z00 | 143 (31%) | 134 (31.5%) | 0.095 |  |  |  |  |  |  |  |  |
| Z01 | 141 (31%) | 82 (19.3%) | <0.001 | MCI (N = 396) |  |  |  |  |  |  |  |
| Z12 | 134 (29.5%) | 74 (17.4%) | <0.001 | ICD | Alive | Deceased | Adj_P_value |  |  |  |  |
| Z13 | 123 (27%) | 67 (15.8%) | <0.001 | F02 | 38 (16.2%) | 5 (3.1%) | 0.002 |  |  |  |  |
| Z78 | 76 (16.7%) | 45 (10.6%) | 0.06 | F03 | 94 (40.2%) | 100 (61.7%) | 0.002 |  |  |  |  |
| Z79 | 173 (38%) | 125 (29.4%) | 0.05 | G31 | 103 (44%) | 48 (29.6%) | 0.09 |  |  |  |  |
| Z98 | 83 (18.2%) | 50 (11.8%) | 0.05 | Z11 | 36 (15.4%) | 4 (2.5%) | 0.002 |  |  |  |  |
|  |  |  |  | Z12 | 66 (28.2%) | 26 (16%) | 0.097 |  |  |  |  |

## Notes

### Competing Interest Statement

The authors have declared no competing interest.

### Funding Statement

This study was funded by National Institutes of Health (NIH) National Institute of Aging (NIA) grant K08AG065519-01A1, UH2AG083254, and the Fineberg Foundation.

### Author Declarations

Ethics committee of University of California, Los Angeles waived ethical approval for this work because all EHRs were deidentified.

